# Quantifying the impact of interventions against *Plasmodium vivax* malaria: a model for country-specific use

**DOI:** 10.1101/2023.02.10.23285652

**Authors:** C. Champagne, M. Gerhards, J. Lana, A. Le Menach, E. Pothin

## Abstract

In order to evaluate the impact of various intervention strategies on *Plasmodium vivax* dynamics, we introduce a simple mathematical model that can be easily adapted to country-specific data. The model includes case management, vector control, mass drug administration and reactive case detection interventions and is implemented in both deterministic and stochastic frameworks. It is available as an R package to enable users to calibrate and simulate it with their own data. By simulating and comparing the impact of various intervention combinations on malaria risk and burden, this model can a useful tool for strategic planning, implementation and resource mobilization.

## 1 Introduction

*Plasmodium vivax* malaria is a parasitic infection responsible for about 4.9 million cases in 2021 [World Health Organization, 2022b]. While generally considered as less severe than *Plasmodium falciparum* malaria, *P. vivax* represents the majority of the remaining cases in many countries where elimination goals have been set for 2025 or 2030, including many countries in Asia (Bhutan, Nepal, Thailand, Republic of Korea, Democratic People’s Republic of Korea), the Pacific (Vanuatu) and the Americas (Costa Rica, Ecuador, Guatemala, Honduras, Mexico, Panama, Suriname) [World Health Organization, 2022b, 2021]. Due to various distinguishing biological characteristics such as its capacity to remain dormant in the liver of infected individuals before its reactivation or its very early transmissibility potential, *P. vivax* is particularly difficult to eliminate. Therefore, reaching the elimination targets requires specific strategies that combine the various available interventions. These interventions include anti-malarial treatments, chemoprevention, case detection and vector control [World Health Organization et al., 2015a, Bassat et al., 2016] and are described in Table 1, highlighting their specific strengths and weaknesses.

**Table 1:** Description of the considered interventions

| Intervention and Mode of action | Advantages | Limitations |
| --- | --- | --- |
| **Treatment/Case management:**<br>• Clearance of blood-stage parasites responsible for the acute infection<br>• Clearance of dormant liver-stage parasites (radical cure) with 8-aminoquinolines such as primaquine (PQ) and tafenoquine (TQ) | • Reduces disease burden [Chu and White, 2021]<br>• Reduces the parasite’s capacity for ongoing transmission by reducing the number of relapses and the number of infectious days [Chu and White, 2021] | • 8-aminoquinolines have restricted eligibility criteria, long treatment scheme and potential adverse effects [Bassat et al., 2016, Chu and White, 2021, Thriemer et al., 2021]<br>• Reaching high effective treatment coverage depends on various components, including physical accessibility to diagnostic and treatment, compliance of the health system or adherence to the treatment scheme [Galactionova et al., 2015, Emerson et al., 2022]<br>• The effectiveness on transmission is limited given the high proportion of patients presenting to health facilities with gametocytes [Douglas et al., 2010] indicating they might already have participated in transmission before being treated. |
| **MDA:** Mass drug administration or mass chemoprevention | • A potential elimination accelerator permitting a rapid reduction of the parasite reservoir in the host population<br>• Deployed in the past in various settings with various success rates [Poirot et al., 2013] | • The evidence from randomized control trials points to a positive short term effect that is most likely not sustained in the longer term if elimination is not reached or if not complemented with additional interventions used in combination [Poirot et al., 2013, Shah et al., 2021].<br>• Logistical, safety and acceptability challenges, especially regarding the mass use of 8-aminoquinolines<br>• Mass use of 8-aminoquinolines not recommended by the World Health Organization [World Health Organization et al., 2015a, Alonso, 2020, World Health Organization, 2022a] |
| **Reactive case detection:**<br>Typically, investigations are conducted in the vicinity of index cases so to enhance the likelihood of finding additional infections that would otherwise have escaped the health system | • Reduces community transmission by identifying infected individuals that have yet to (and may not plan to) seek care<br>• Part of the WHO strategic pillar to “transform malaria surveillance into a core intervention” [World Health Organization et al., 2015b]<br>• Credited to contributing to the successful elimination strategies in China and El Salvador [Perera et al., 2020, Burton et al., 2018, Balakrishnan, 2021] | • Challenges in the feasibility of this resource intensive activity which may take resources away from routine surveillance and diagnosis activities<br>• Uncertainty in the conditions for its effectiveness and difficulty to monitor effectiveness [van Eijk et al., 2016, Perera et al., 2020, Hetzel and Chitnis, 2020] |
| **Vector control:** Killing or repelling the *Anopheles* mosquitoes responsible for parasite transmissions, via interventions such as insecticide treated bednets (ITNs) or indoor residual spraying (IRS) | • Complementary elimination tool to reduce the likelihood of transmission when cases are not treated promptly or correctly<br>• Key component of the WHO recommended prevention strategies [World Health Organization et al., 2015a]<br>• ITNs has been one of the most important drivers of the reductions in malaria burden in Africa in the beginning of the 21st century [Bhatt et al., 2015] | • Many eliminations settings outside of Africa harbour mosquitoes species with different characteristics, such as early biting behaviours or different response to insecticides that reduce intervention effectiveness [Briët et al., 2019, Monroe et al., 2020].<br>• Logistical challenges to optimize deployment timing given the limited duration of effectiveness (especially IRS)<br>• Acceptability and logistical challenges to reaching high coverage/usage of the interventions<br>• Short term durability of the insecticides and nets, requiring frequent re-deployment of the interventions |

Because of the advantages and challenges involved with each type of intervention, defining the most appropriate combination of interventions highly depends on context-specific factors, such as the level of transmission, ecological factors, the vulnerability to case importation or the characteristics of the health system [Cohen et al., 2017]. Therefore, the choice and adaptation of the malaria intervention strategy required to reach elimination targets needs to be tailored to local context. Mathematical modelling can be used to quantify the impact of the considered intervention strategies, identify the most impactful ones in each setting [Owen et al., 2022]. Various *P. vivax* models have recently been developed (e.g. [White et al., 2018, Kim et al., 2021, Tian et al., 2022]), but they are not always readily operationalized for being used routinely at country level, either because they don’t include all the above-mentioned interventions, or because their calibration to routine data is not straightforward to implement.

In order to address these shortcomings, a model was previously developed to represent *P. vivax* dynamics at the local level in the presence of case management interventions, including a methodology to infer parameter values from available data based on the steady-state assumption [Champagne et al., 2022]. Nonetheless, this model has some limitations that restrict its use in practice. Firstly, the model assumes that treatment acts instantaneously, such that treated patients do not contribute to ongoing disease transmission [Douglas et al., 2010]. This assumption is not totally appropriate for *P. vivax*, for which treated patients might already have participated in transmission before the effect of their treatment. Secondly, it does not include other interventions such as reactive case detection (RCD) or mass drug administration (MDA), which are part of the malaria elimination toolbox for decision-makers. Finally, it is only implemented in the deterministic framework.

The present work therefore extends the model by Champagne et al. [2022] by removing these limitations, thus increasing its potential applications for country-specific decision making. Such use-cases are illustrated in an example on three fictitious areas of varying endemicity. The extended model is publicly available as an R package (https://swisstph.github.io/VivaxModelR/) that can be applied to the users’ own data.

## 2 Methods

### 2.1 Model of *P. vivax* dynamics with delayed treatment

*P. vivax* dynamics are represented by a compartmental model where the host population is divided between infectious and susceptible individuals who do or do not harbour liver stage parasites [White et al., 2016, Champagne et al., 2022]. In order to allow the possibility for treated individuals to infect mosquitoes before they effectively clear their parasites, the model by Champagne et al. [2022] is modified by adding two compartments representing treated individuals. Let *T*_*L*_ be the proportion of blood-stage infected population with liver-stage infection which received treatment, *T*_0_ that of blood-stage infected population without liver-stage infection which received treatment, *U*_*L*_ that of blood-stage infected population with liver-stage infection but which did not received treatment and *U*_0_ that of blood-stage infected population without liver-stage infection which did not receive treatment. We define *I* := *T*_*L*_ + *T*_0_ + *U*_*L*_ + *U*_0_ as the proportion of blood-stage infections. *S*_0_ represents the proportion of fully susceptible individuals, and *S*_*L*_ represents the individuals who have cleared their blood stage parasites but still harbour liver stage parasites and hence have the possibility to experience a relapse. The model can be represented by the following system of equations:

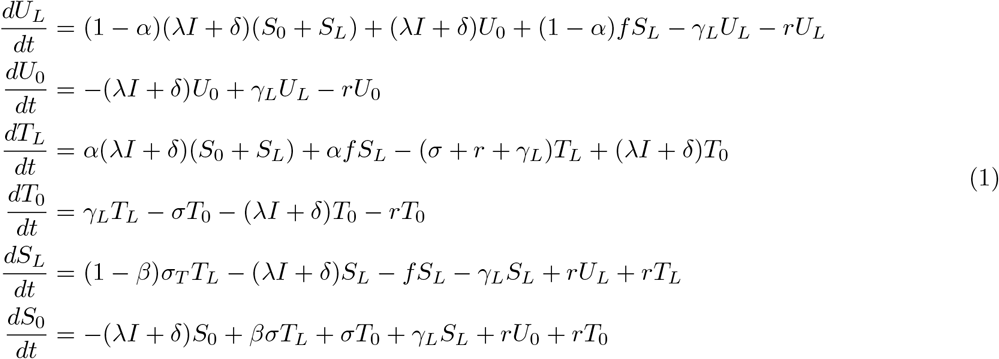

All other parameter notations are indicated in Table 2 and a schematic representation of the model is presented in Figure 1. In this model, case management is thus represented via three parameters. The first parameter is the proportion of individuals receiving effective treatment (*α*), i.e. the proportion of individuals whose blood-stage parasites are effectively cleared due to treatment, if they don’t recover naturally before. The second parameter (*β*) is the proportion of treated individuals who experience radical cure, i.e. the clearance of liver-stage parasites in addition to blood-stage parasites. The third parameter (*σ*) quantifies the time during which a treated individual can transmit the disease to mosquitoes (ignoring potential recoveries that could happen during this interval, see Appendix A for more details). All three parameters can be informed by data on the health system, following the effective coverage framework [Galactionova et al., 2015]. A description of the rationale for choosing the formulation in equation (1) is presented in Appendix A in the context of perfect radical cure.

**Table 2:** Description of state variables and model parameters.

| Notation | Description | Unit | Definition range |
| --- | --- | --- | --- |
| **State variables** |  |  |  |
| $U_L$ | Untreated individuals with liver and blood stage parasites | . | [0,1] |
| $U_0$ | Untreated individuals with blood stage parasites only | . | [0,1] |
| $S_L$ | Susceptible individuals with liver stage parasites | . | [0,1] |
| $S_0$ | Fully susceptible individuals | . | [0,1] |
| $T_L$ | Treated individuals with liver and blood stage parasites | . | [0,1] |
| $T_0$ | Treated individuals with blood stage parasites only | . | [0,1] |
| **Parameters** |  |  |  |
| $\lambda$ | Transmission rate | time<sup>-1</sup> | $\geq 0$ |
| $r$ | Blood stage clearance rate | time<sup>-1</sup> | $\geq 0$ |
| $\gamma_L$ | Liver stage clearance rate | time<sup>-1</sup> | $\geq 0$ |
| $f$ | Relapse frequency | time<sup>-1</sup> | $\geq 0$ |
| $\delta$ | Importation rate | time<sup>-1</sup> | $\geq 0$ |
| $\alpha$ | Proportion of effective treatment | . | [0,1] |
| $\beta$ | Proportion of radical cure | . | [0,1] |
| $\sigma$ | Duration of infectivity for treated infections | time<sup>-1</sup> | $\geq 0$ |
| $\rho$ | Observation rate | . | [0,1] |
| **RCD model** |  |  |  |
| $\iota_{max}$ | Maximal number of index cases investigated | time<sup>-1</sup> | $\geq 0$ |
| $\nu$ | Number of individuals investigated per index case | . | $\geq 0$ |
| $\eta$ | Proportion of effective care for infections detected by RCD | . | [0,1] |
| $\tau$ | Targeting ratio | . | $\geq 0$ |
| **MDA model** |  |  |  |
| $c_{MDA}$ | MDA coverage | . | [0,1] |
| $\beta_{MDA}$ | Proportion of radical cure for MDA | . | [0,1] |
| $t_{MDA}$ | Starting date of MDA | time | $\geq 0$ |
| $p_{MDA}$ | Duration of MDA prophylaxis | time | $\geq 0$ |

**Figure 1:**
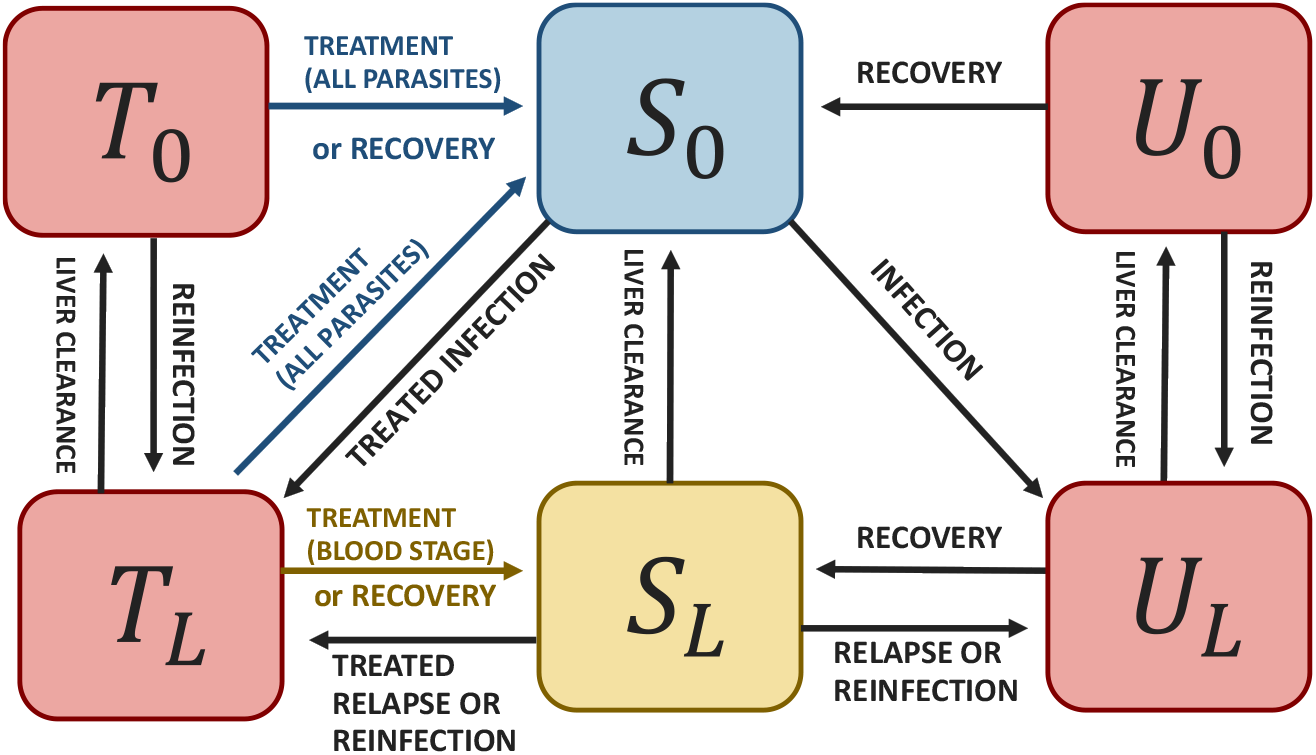
Schematic representation of the model for *P. vivax* dynamics including delayed access to treatment.

The corresponding reproduction numbers in the presence of control interventions (*R*_*c*_) and in the absence of control (*R*_0_) are calculated using the next-generation matrix approach [van den Driessche and Watmough, 2002] as follows:

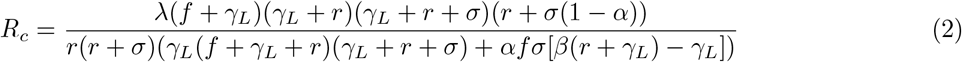

and

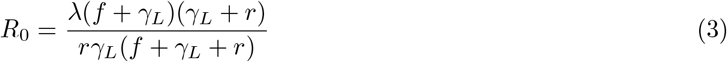

As expected, the basic reproduction number *R*_0_ in the absence of control is identical to the one in the model where the effect of delay in treatment access was neglected [Champagne et al., 2022], because in this hypothetical context no individual receives any treatment. Similarly to Champagne et al. [2022], a polynomial relationship between the transmission rate *λ* and observable quantities can be calculated, as detailed in Appendix B. With this relationship, the model can easily be calibrated to reported incidence data.

### 2.2 Including reactive case detection

Reactive case detection (RCD) can be included in the model based on the framework by Chitnis et al. [2019]. With this approach, reactive case detection adds the possibility for non-treated cases to be detected and treated. The model relies on the assumption that cases are geographically clustered, such that the probability of finding a case in the vicinity of a reported case is higher than the probability of finding a case in the general population. This increased likeliness of finding cases is modelled via the targeting ratio *τ* as in Chitnis et al. [2019], Das et al. [2022a]. The parameter *ι* indicates the number of index cases investigated per population per unit of time. The parameter *ν* indicates the number of secondary individuals investigated per index case. We add the parameter *η* to reflect the effective cure of RCD-detected individuals (including test sensitivity, compliance, adherence and drug efficacy). A schematic representation of the model is presented in Figure 2. Cases detected via RCD are assumed to receive effective radical cure with the same probability as other treated infections.

**Figure 2:**
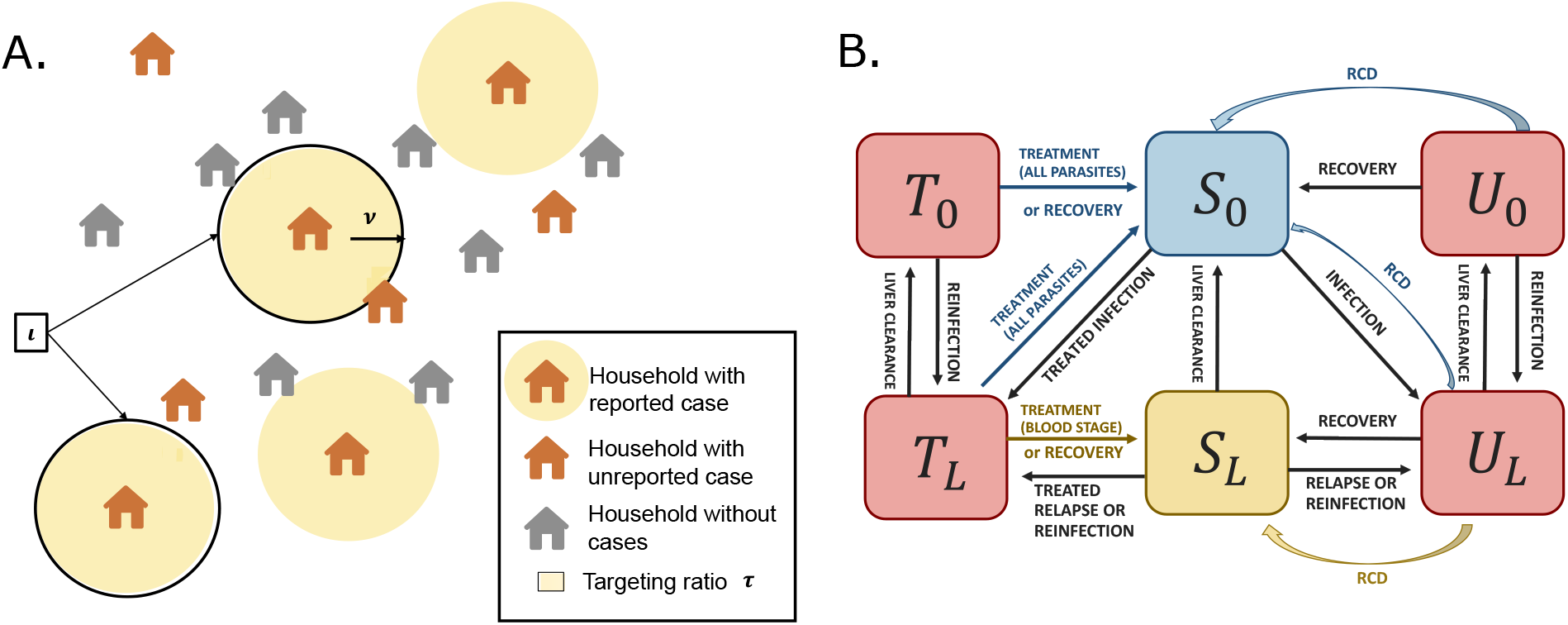
Models including reactive case detection (RCD). A. Description of the assumptions for modelling RCD. B. Schematic representation of the model for *P. vivax* including delayed treatment and RCD (another model version in which cases detected via RCD need to be referred to a health facility for treatment and would experience a delay before being cured is presented in Appendix C.3).

The model with delay in treatment and reactive case detection, corresponding to Figure 2B, can be represented by the following equations:

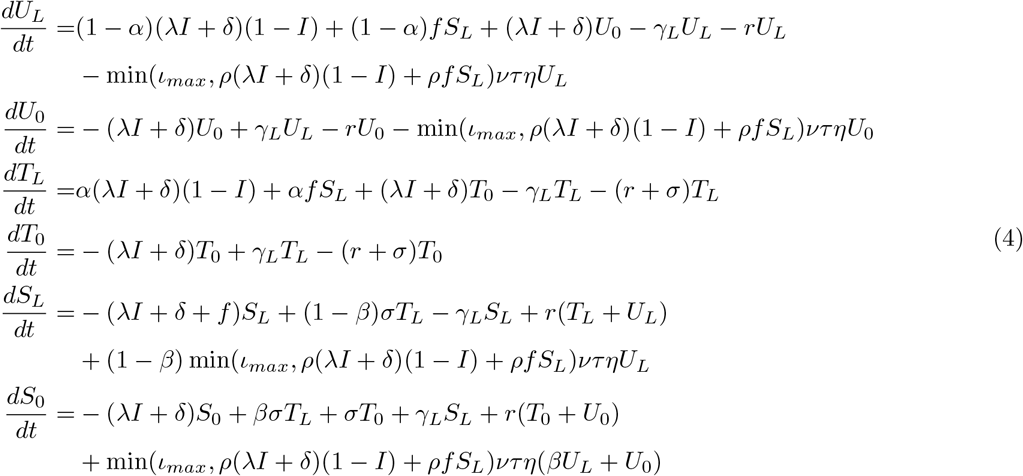

The number of index cases investigated is written as *ι* = min(*ι*_*max*_, *ρ*(*λI* + *δ*)(1 − *I*) + *ρf S*_*L*_) and this formulation ensures that the number of index cases investigated is never higher than the number of cases detected.

This model makes the assumption that cases detected by RCD are immediately effectively treated (transition from *U*_*L*_ or *U*_0_ to *S*_*L*_ or *S*_0_). In some situations, the cases detected via RCD need to be referred to a health facility for treatment, hence they would experience a delay before being cured. Therefore, another version of the model in which infections detected via RCD enter the *T*_0_ and *T*_*L*_ compartments instead of *S*_0_ and *S*_*L*_ was also developped and is presented in Appendix C.3 (called “with referral to health facility”). For completeness, the model with RCD but without delayed access to treatment is also presented in Appendix C.1.

If *ιντη* = 0, this model reduces to the model (1) without RCD. If *τ* is fixed (or at least bounded), all of the ‘RCD terms’ are of the order *O*(*I*^2^) for *I* → 0, so the reproduction number *R*_*C*_ is equal to *R*_*C*_ in the model without any RCD. This means that RCD as modelled cannot interrupt sustained disease transmission, nor can it overcome the effect of importation as soon as local prevalence is too low. So either with or without importation, RCD can only affect the magnitude of the endemic equilibrium and not the threshold behaviour between endemic and disease-free equilibrium. The intuition behind this phenomenon is that, as prevalence decreases, both the number of detected index cases and the number of cases found in each investigation decrease, such that the number of cases found by RCD is reduced. In practice, it could happen that this effect is compensated by another one, such as an increased clustering of cases around index cases represented with unbounded targeting ratio *τ*. For example, following Chitnis et al. [2019], *τ* could also be a time-varying quantity, which depends on the prevalence in the population and the number of secondary cases investigated, such that the targeting ratio increases as prevalence decreases and as *ν* decreases. A parametric function of the prevalence and *ν* fitted to data from Zambia is presented in Chitnis et al. [2019] and further used in Reiker et al. [2019] and could be substituted to the fixed values of *τ*.

In all cases, extinction events may also occur when the endemic equilibrium is very low and the model is simulated in the stochastic framework (cf. below).

#### 2.2.1 Relation between the transmission rate and observable quantities

The transmission rate *λ* can be calculated from observable quantities using the model’s equilibrium with a similar methodology to Champagne et al. [2022], where a polynomial equation for *λ* is derived and solved numerically. The extension of this framework to the models with RCD is detailed in Appendix C. Nonetheless, in order to use these polynomial equations in practice for calculating *λ*, additional calculations to derive *I*\*, *δ* and *ι*\* from observable quantities are required. Let us introduce the following additional notations:

- at equilibrium, we note 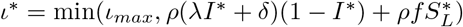
- 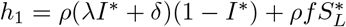 is the incidence of directly-detected infections
- 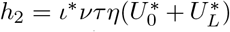 is the incidence of reactively-detected infections (assuming they are all perfectly reported).
- *h* = *h*_1_ + *h*_2_ is the total incidence of detected cases
- *p* is the proportion of imported cases, such that *ph* = *ρδ*(1 − *I*\*)

With these notations, 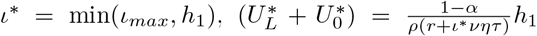 (using (28) in Appendix C), 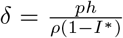 and *I*\* can be calculated using (32) in Appendix C. Therefore, one needs *h*_1_ to back-calculate the value of *λ*.

Two situations can arise in practice:

1. only the total number of new cases *h* is known, regardless of whether cases are detected via direct detection or via reactive detection
2. the numbers of reactively detected and non-reactively detected cases are both known (*h*_1_ and *h*_2_).

#### 2.2.2 Only the total number of new cases is known

In the first situation, we need to calculate the value of *h*_1_, and the value of *h*_2_ will be *h*_2_ = *h* − *h*_1_.

We need to evaluate separately the two possibilities for *ι*.

If *ι* = *ι*_*max*_, we have 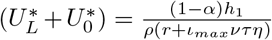. Hence 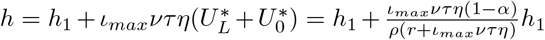 and

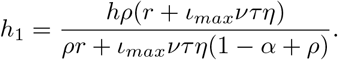

If *ι* = *h*_1_, we have 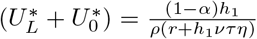. Then, from 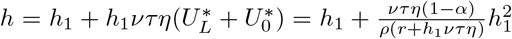, we get

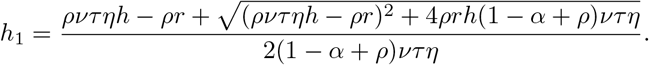

Combining these two results, in the case of capped *ι*, we find

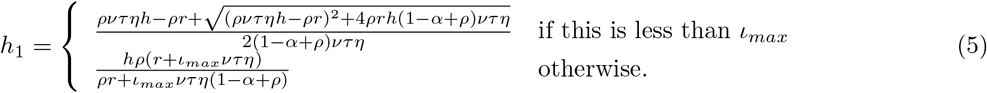

#### 2.2.3 The numbers of reactively detected and non-reactively detected cases are both known

If we know both *h*_1_ and *h*_2_, we can calculate *ι*\*, *I*\*, *δ* and therefore *λ* directly. Additionally, we can combine this information to get an estimate of *τ*, which is otherwise difficult to find. We will consider only the cases where *h*_2_ > 0 (otherwise, there is no effect of RCD and therefore no reason for using *h*_2_ to calculate *τ*

From equation (28) in Appendix C, we can infer

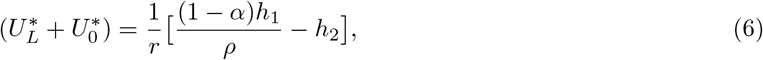

which can be used in

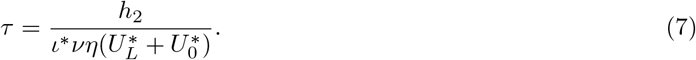

### 2.3 Including Mass Drug Administration (MDA)

In order to model the deployment of an MDA campaign, the state variables representing the infectious population are depleted at the time of the MDA deployment depending on the MDA coverage. In order to model the time during which drug prophylaxis prevents targeted individuals from reinfections following MDA deployment, another model including two additional compartments is used. Finally, at the end of the prophylaxis time, the initial model without MDA can be simulated from the newly found initial condition. The overall framework is presented graphically in Figure 3.

**Figure 3:**
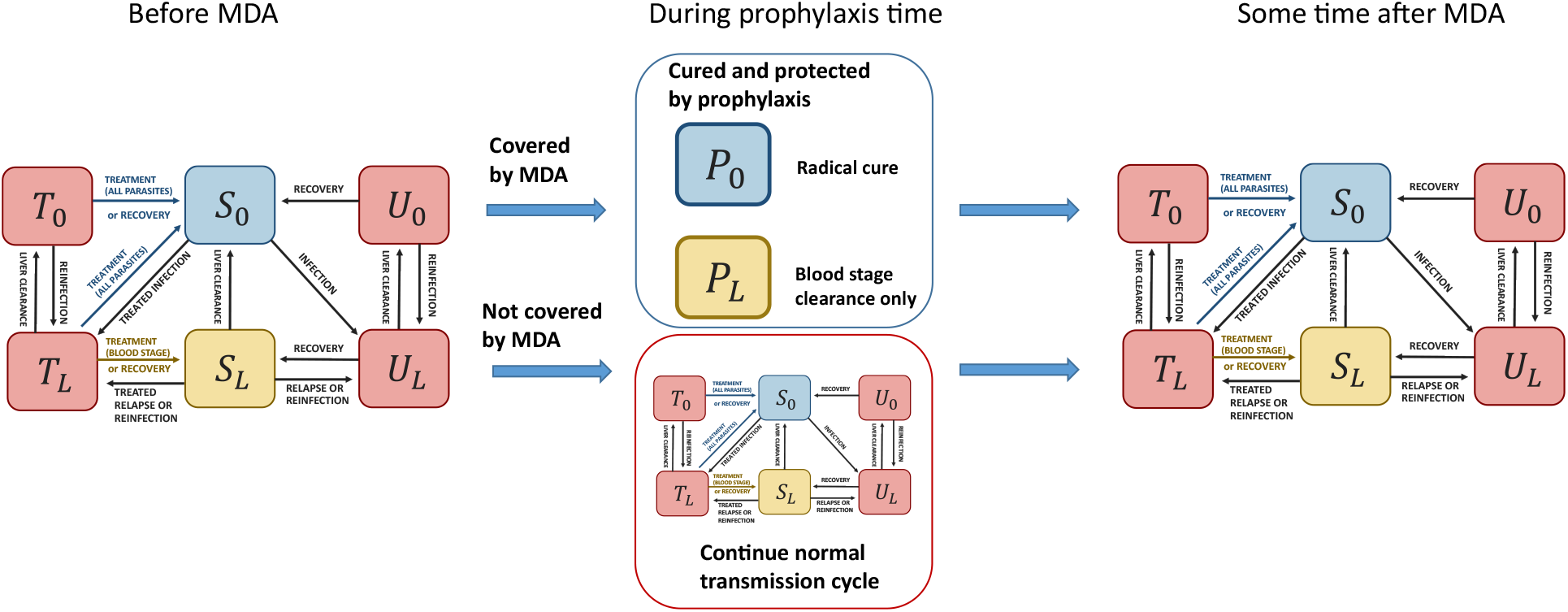
Models including MDA (example of the model with delayed treatment).

We present here the equations of the model with delayed treatment and without RCD (1) as an example. Other model options with RCD or without delayed access to treatment follow the same approach and are detailed in Appendix D.

The simulation starts with the models defined by (1). The parameters of MDA, namely effective coverage *c*_*MDA*_, starting time of prophylaxis *t*_*MDA*_, duration of prophylaxis *p*_*MDA*_ and percentage of radical cure *β*_*MDA*_, do not come into play in the ODEs. Instead, they affect the state variables of the system at a fixed point in time. At the time of MDA deployment *t*_*MDA*_, the state variables of the system are modified as follows (using the notation 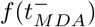 for the limit of *f* (*t*) as *t* appproaches *t*_*MDA*_):

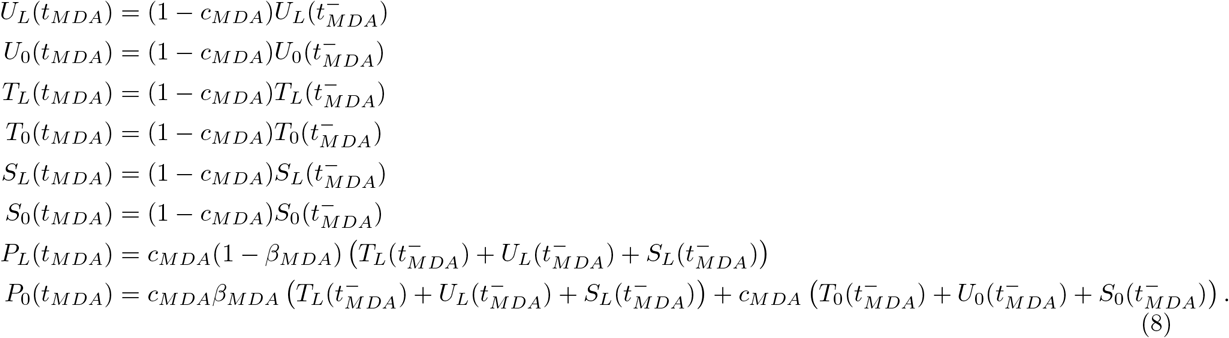

To model the two effects of MDA (treatment and prophylaxis), we include two new compartments in the model. Let *P*_*L*_ and *P*_0_ be the proportions of people that are effectively covered by MDA with and without liver-stage infection, respectively. We have the following set of ODEs to describe the dynamics of the system during the prophylaxis period:

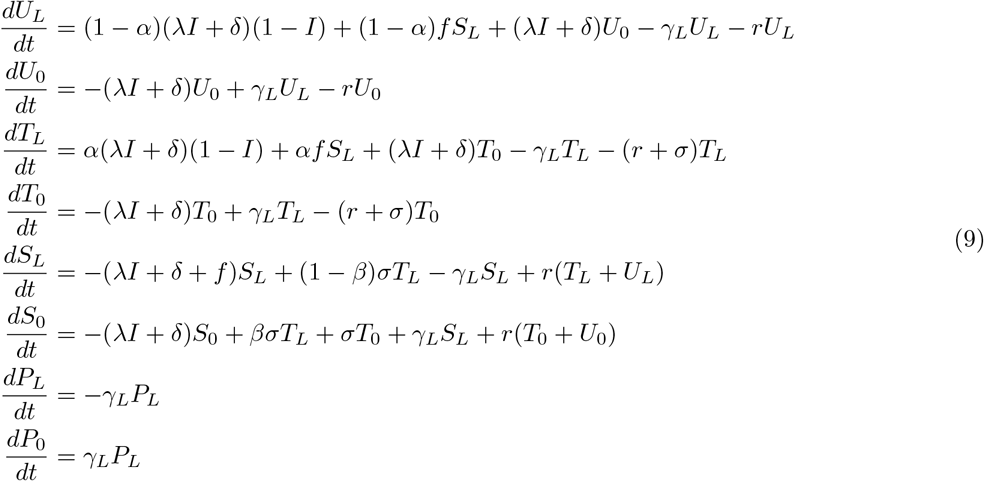

starting with the initial conditions defined in (8).

We are assuming constant level of prophylaxis in (9). Each individual either has perfect prophylaxis during the intervention or none at all in which case they go through the usual infection pathway.

Finally, at time *t*_*MDA*_ + *p*_*MDA*_, the state variables are updated as follows:

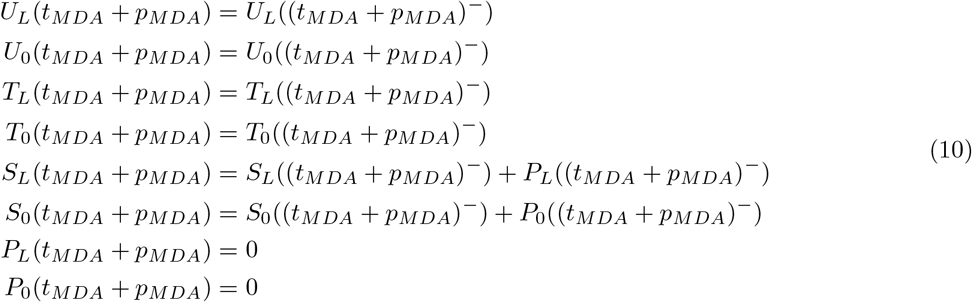

and the initial model (1) is further simulated from these initial conditions.

Several MDA rounds can be chained using the same approach.

### 2.4 Including vector control

As mentioned in Champagne et al. [2022], vector control can be included in the model as a reduction of the intensity of transmission *λ*, following Briët et al. [2019]. The transmission parameter *λ* becomes *ωλ* where *ω* ∈ [0, 1] represents the intensity of vector control and can be informed by an external model for vector dynamics [Briët et al., 2019, Golumbeanu et al., 2022]. The absence of vector control corresponds to the case *ω* = 1, and *ω* = 0 represents perfect vector control that completely disables vector-borne transmission. By making *ω* time-varying, one can include a decay in vector control effectiveness, for example due to the waning of insecticide or the attrition of bednets, as in [Briët et al., 2019, Briet et al., 2020, Golumbeanu et al., 2022].

### 2.5 Model implementation

The model is implemented in R [R Core Team, 2019] as a publicly available package (https://swisstph.github.io/VivaxModelR/). The package enables the user to calibrate and simulate all the model versions presented here (with and without delays in treatment, with and without RCD, with and without MDA). The package includes tests to ensure that the back-calculations and simulations with the various models are correctly implemented, as well as a user tutorial.

The ordinary differential equations solving relies on the *deSolve* package [Soetaert et al., 2021] and a stochastic implementation with either Gillespie algorithm or *τ* -leap methodologies is provided relying on the *TiPS* package [Danesh et al., 2020, 2021]. Thanks to the convergence of the stochastic version of the model to its ODE counterpart for infinite population sizes [Kurtz, 1970], the back-calculation of the transmission rate obtained with the ODE steady-state can be used in the stochastic model and the coherence between the two model implementations is guaranteed. Such convergence is empirically verified as part of the set of automated tests included in the R package.

When simulating future intervention scenarios with the model, all implementations offer the possibility for the importation rate *δ* and the vector control term *ω* to be time-varying, using any empirical function provided by the user as a dataset.

## 3 Application: an illustrative example

As an illustrative example, we simulate three fictitious areas with varying reported case numbers, assuming perennial *P. vivax* transmission and values similar to Champagne et al. [2022], as indicated in Table 3. The number of reported cases in each area increases from very low in Area 1 to moderate-high in Area 3. In

**Table 3:** Model parameters and data at baseline for three fictitious areas.

| Baseline parameters | Area 1 | Area 2 | Area 3 |
| --- | --- | --- | --- |
| **Setting-specific data** |  |  |  |
| Reported cases per year | 6 | 95 | 540 |
| Imported cases per year | 1 | 0 | 27 |
| Population size | 5000 | 5000 | 5000 |
| **Setting-specific parameters at baseline** |  |  |  |
| Access to care ($a$) | 0.5 | 0.5 | 0.5 |
| Proportion of infections effectively cured for blood stage ($\alpha$) | $a * s * d$ | $a * s * d$ | $a * s * d$ |
| Radical cure ($\beta$) | $0.86 * \epsilon$ | $0.86 * \epsilon$ | $0.86 * \epsilon$ |
| Observation rate ($\rho$) | $\alpha$ | $\alpha$ | $\alpha$ |
| Average treatment delay ($1/\sigma$, in days) | 10 | 10 | 10 |
| Vector control ($\omega$, $\omega = 1$ corresponds to the absence of control) | 1 | 1 | 1 |
| RCD: max. index cases investigated ($\iota$, per week per 5000 inh.) | 50 | 50 | 50 |
| RCD: number of individuals investigated per index case ($\nu$) | 5 | 5 | 5 |
| RCD: targeting ratio ($\tau$) | 5 | 5 | 5 |
| RCD: probability to detect and treat a case ($\eta$) | $d$ | $d$ | $d$ |
| MDA: coverage ($c_{MDA}$) | 0 | 0 | 0 |
| MDA: radical cure treatment ($\beta_{MDA}$) | 0 | 0 | 0 |
| **Fixed parameters** |  |  |  |
| Relapse frequency ($f$, in days<sup>-1</sup>) | 1/223 [White et al., 2016] | | |
| Clearance rate ($r$, in days<sup>-1</sup>) | 1/60 [White et al., 2016] | | |
| PQ efficacy ($\epsilon$) | 0.76 [Champagne et al., 2022] | | |
| RDT sensitivity ($d$) | 0.95 [Abba et al., 2014] | | |
| Symptomatic proportion ($s$) | 0.3 [Cheng et al., 2015] | | |
| **Estimated reproduction numbers** |  |  |  |
| $R_0$ | 0.79 | 1.38 | 1.39 |
| $R_c$ | 0.64 | 1.11 | 1.12 |

Area 2, it is assumed that there is no importation and all cases are contracted locally. The three areas are assumed to have the same intervention parameters at baseline for case management, reactive case detection and vector control. MDA being a transient intervention, it is assumed to be absent at baseline.

When calibrating the model at baseline with the chosen parameter values, malaria is sustained through importation in Area 1 as *R*_*c*_ is smaller than 1 (cf. Table 3). On the contrary, Areas 2 and 3 experience sustained local transmission (*R*_*c*_ > 1), a result that was expected by construction in Area 2 due to the assumed absence of importation.

From there, five intervention scenarios are simulated, as detailed in Table 4. The first three interventions are case management strengthening, reactive case detection strengthening and deployment of indoor residual spraying (IRS): the parameters corresponding to these interventions are detailed in Table 4 and their impact on malaria prevalence is presented in Figure 4. In this example, case management strengthening always leads to a large decrease in malaria prevalence although it is not sufficient to reach elimination. Reactive case detection has a much larger impact in areas with higher endemicity as there are more cases to be detected. In the three examples considered, using the time-varying targeting ratio from Chitnis et al. [2019] leads to stronger prevalence reductions than the fixed value, as the targeting ratio increases with decreasing prevalence and hence accelerate the decrease in malaria trends. Finally, IRS leads to important reductions in malaria prevalence, but because its 6-month effectiveness duration, the intervention needs to be redeployed regularly to sustain the gains. In this particular example, improvements in case managements have a greater impact on prevalence compared to the introduction of IRS or the strengthening of RCD.

**Table 4:**
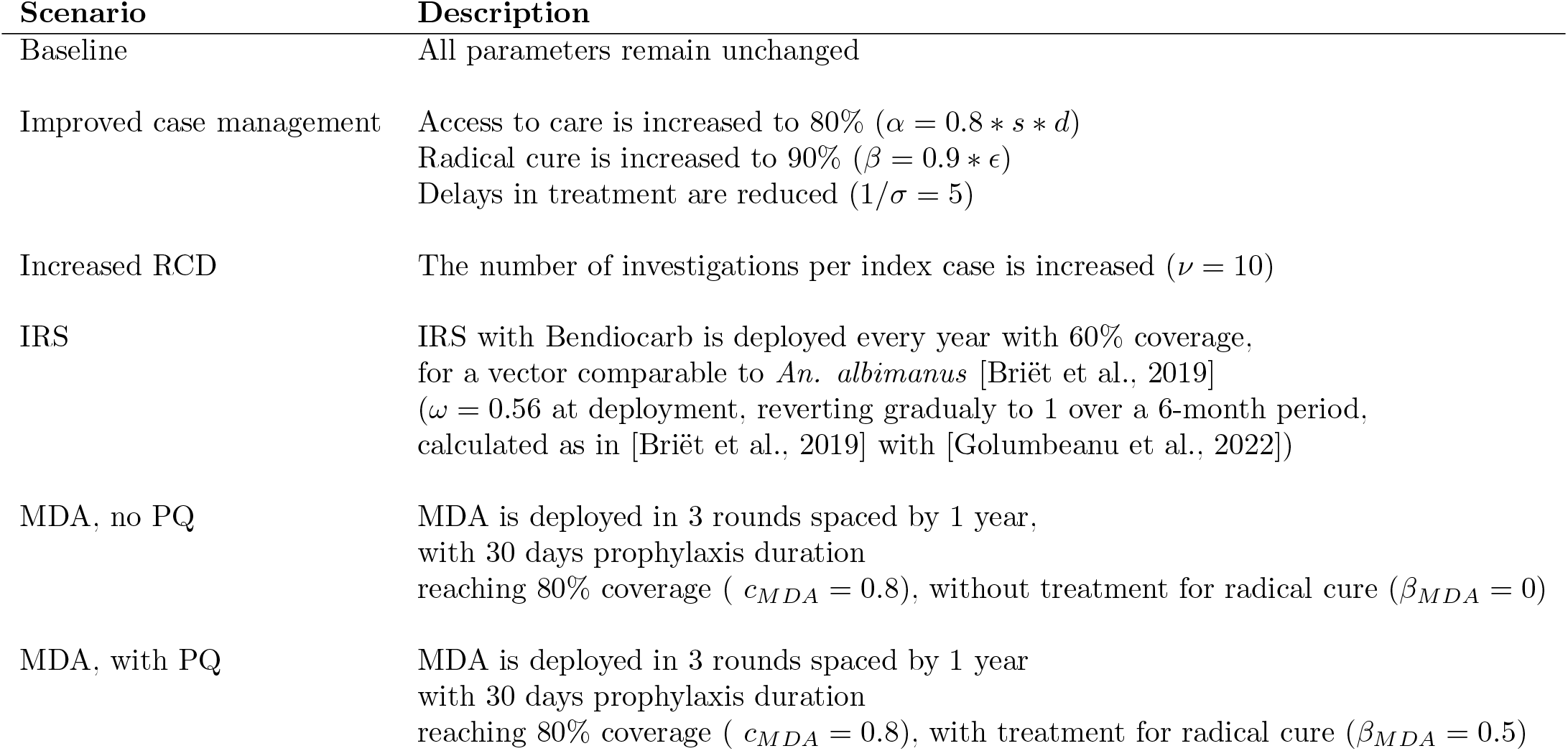
Parameter values of the intervention scenarios simulated

**Figure 4:**
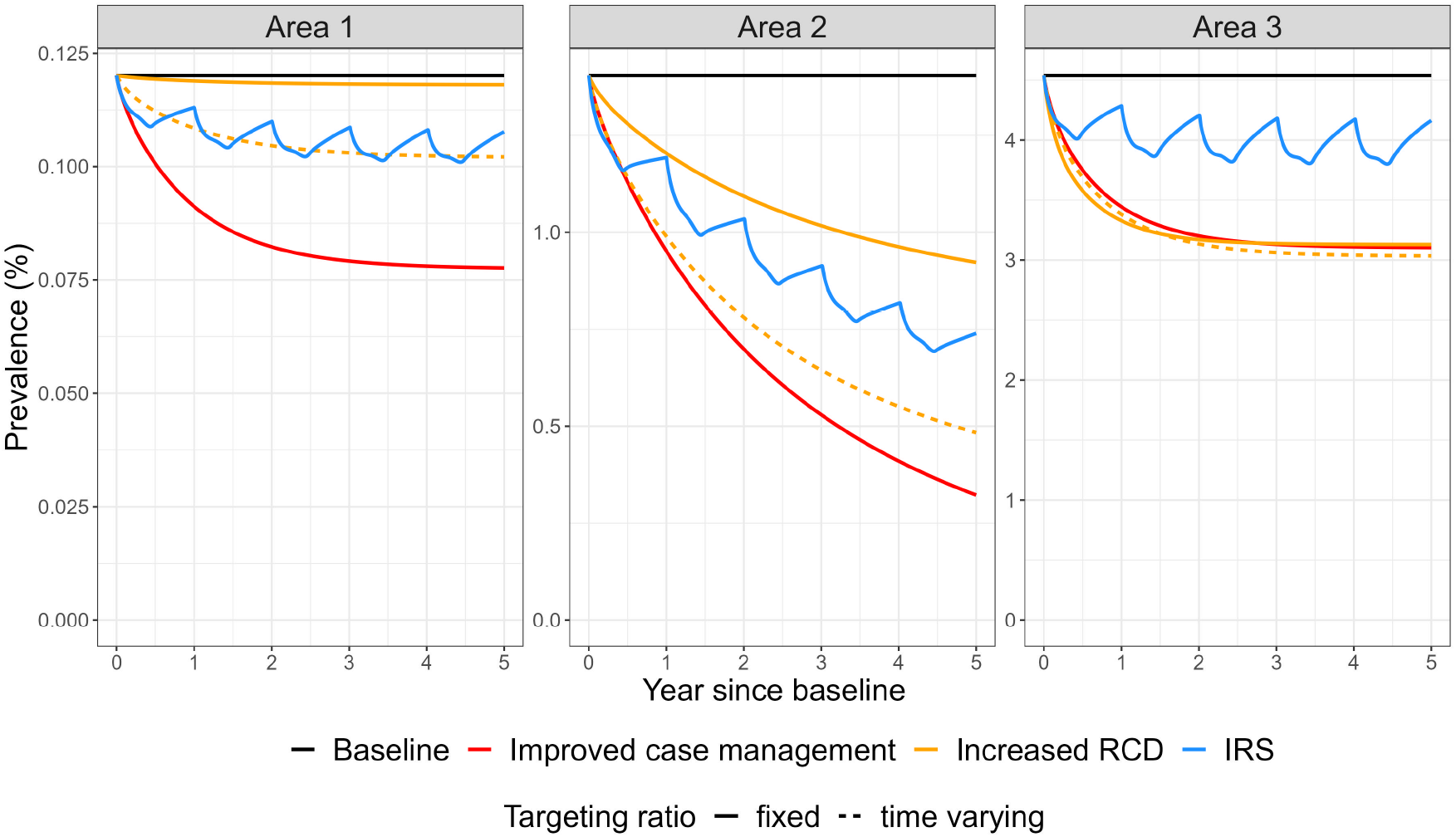
Effect of various intervention scenarios on *P. vivax* prevalence, applied in three artificial settings. The parameter values corresponding to each scenario can be found in Table 4.

**Figure 5:**
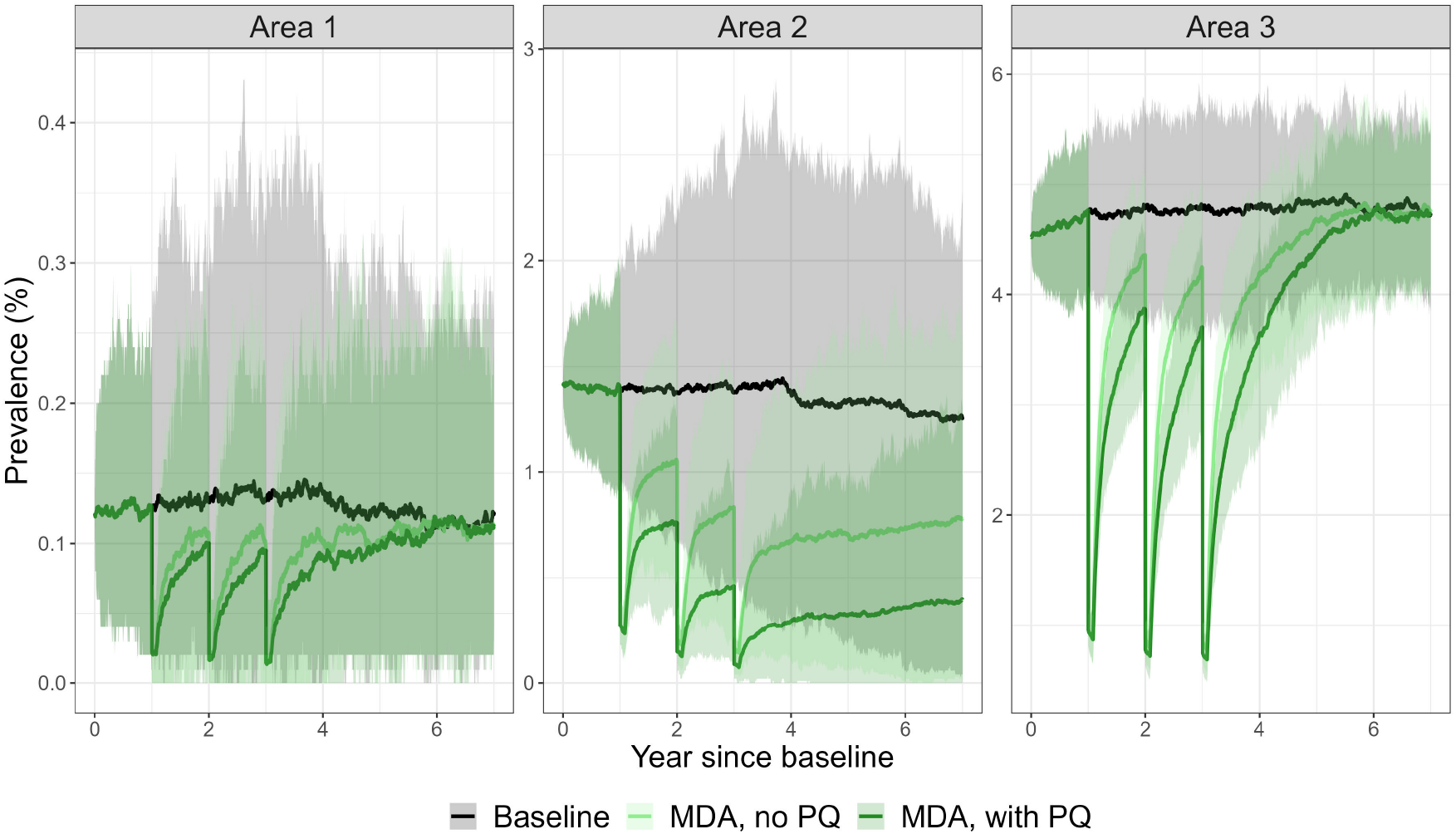
Effect of MDA scenarios on *P. vivax* prevalence, applied in three artificial settings. The parameter values corresponding to each scenario can be found in Table 4. The lines represent the median of 100 independent stochastic simulations, while the shaded area represent the 95% uncertainty intervals.

The last two scenarios include MDA with and without PQ for radical cure. Because of the high variability expected when the model reaches low case numbers, the stochastic version of the model is used, with 100 independent simulation replicates. The effect of MDA is transient, and malaria transmission bounces back to initial prevalence levels when the intervention is interrupted. As expected, targeting the liver-stage reservoir with PQ increases the prevalence reduction achieved. In the scenario with very low transmission (Area 1), elimination is not reached within the 7 years simulated because transmission is sustained with importation, elimination being defined in this example as a whole year with zero cases (including reported and unreported). Other definitions of malaria elimination in the presence of importation, distinguishing local, indigenous and imported cases [Das et al., 2022b] could also be explored but are beyond the scope of the current work. In the scenario without importation (Area 2) and with PQ MDA, elimination in 7 years is reached in 11% of the simulations, highlighting the difficulty of reaching this outcome.

## 4 Discussion

In this work, we presented an extended compartmental model for *P. vivax* dynamics at the population level that includes four commonly deployed interventions, namely treatments, vector control, reactive case detection and mass drug administration. It can be calibrated in each setting of interest at the local level using the observed incidence at steady-state, thus providing estimates of transmission potential. The setting-specific model can then be used to compare the impact of various intervention strategies and identify the most promising ones. The model can be simulated in both the deterministic and the stochastic framework and it is readily implemented as a publicly available R package.

Thanks to its stochastic implementation [Danesh et al., 2020, 2021], the model can be used to compute elimination probabilities and timelines, which are useful outcomes to compare malaria intervention strategies in low-endemicity settings. Additionally, the inclusion of demographic stochasticity opens the possibility to use the model for small population sizes, as required to represent *P. vivax* dynamics at local scales.

The model incorporates the effect of routine malaria treatment on transmission through three parameters: the proportion of blood-stage infections that are treated, the proportion of treated cases that achieve radical cure of the liver-stage parasites and the duration of infectivity for treated cases. These three aspects are important components of health system strengthening strategies and the possibility to compare improvements in these three dimensions simultaneously or independently can provide useful information for decision-makers. Nonetheless, it is important to note that the model is very sensitive to the value of baseline case management parameters [Champagne et al., 2022] and the modelled impact of case management strategies relies heavily on the availability and quality of data on the health system at the spatial scale of interest.

The mathematical representation of RCD relies on the definition of a targeting ratio, and thus does not include mechanistically the effect of case clustering as would be the case in spatially-explicit individual-based models [Gerardin et al., 2017]. Therefore, similarly to Chitnis et al. [2019], Reiker et al. [2019], Das et al. [2022a], it is necessary to parameterise the targeting ratio using setting-specific data [Das et al., 2022a] or by choosing appropriate assumptions adapted to the local context. Epidemiological knowledge of infection risk factors to evaluate if such a strategy is suitable in a given setting is therefore considered as a prerequisite before using the model. For example, in countries where *P. vivax* infection is mainly driven by occupational exposure (e.g. working in the forest), the investigation of the neighborhood of index cases might be less efficient compared to occupational screening [Mukaka et al., 2021]: this effect could be represented with a smaller targeting ratio. This appreciation is therefore left to the user when choosing the appropriate value for targeting ratio in the setting of interest. Nonetheless, if data on the source of case reporting (via RCD or via other detection channels) is available, the value of the targeting ratio can be quantified at baseline using the back-calculation methodology presented here. The objective of the current model is therefore not to investigate if RCD has the capacity to detect cases in general, but rather to evaluate its impact in relation to other interventions and for various implementation designs.

In line with other modelling work [Robinson et al., 2015, Pemberton-Ross et al., 2017, White et al., 2018, Obadia et al., 2022] and with the available epidemiological evidence [Shah et al., 2021], MDA is represented as an instantaneous shock on the state variables without affecting the model parameters. Therefore, its effect is transient by construction and malaria dynamics are expected to revert to their previous equilibrium except in the stochastic case if elimination is reached, or if accompanied by sustainable changes such as increased intervention coverages or environmental modifications. Decay in the effectiveness of prophylaxis is not explicitly modelled, rather the model assumes a constant level of protection during the time of prophylaxis. This simplification should not strongly affect the annual results for drugs with prophylaxis duration of 15 to 30 days. Nonetheless, the model could also be extended to include parametric decay forms.

Mosquito dynamics are not directly included in the model and the effect of vector control is represented as a reduction of the human-to-human transmission intensity. This simplifying assumption is chosen because the time scale of vector dynamics is much shorter than the one of human dynamics, such that in a low endemicity setting where the proportion of infectious host is small, the parasite dynamics can be approximated by an SIS-like model [Smith and McKenzie, 2004]. Additionally, using the model by Golumbeanu et al. [2022] enables the user to account for differences in vector control efficacy due to mosquito species biological characteristics, intervention type or the proportion of time that individuals spend indoors or in bed [Briët et al., 2019], whose importance can be crucial, especially in elimination settings outside of Africa.

Seasonality is not included in the model, which is suitable for perennial settings or when studying annual data and interventions that do not have seasonal effects. Nonetheless, the R package implementation of the model offers the possibility to include a seasonal forcing into the transmission rate via the time-varying parameter *ω* when simulating disease dynamics over time. Adapting the back-calculation methodology and reproduction number definition to the seasonal case [Bacaër, 2007] is however outside the scope of the current work.

The model is a simplified representation of reality and has therefore a certain number of limitations. Firstly, the chosen representation of treatment delays assumes that host infectiousness to mosquitoes is constant over time, which is an important simplification in regard to the complex life cycle of the malaria parasites [Gaspoz and al., in prep] and the relaxation of this assumption will be the object of future work. In addition, delays in access to treatment were represented with exponential durations via the ODE formalism and not fixed durations as would have been the case with delay differential equations [Kim et al., 2021, Tian et al., 2022]. Nonetheless, the chosen formalism of the model presents some similarities with a model based on delayed-differential equations, as illustrated in Appendix A for a simpler model with perfect radical cure.

Moreover, because of its emphasis on implementation simplicity, this model makes some additional simplifications in the biological depiction of *P. vivax*. Importantly, the model does not include any form of immunity, an assumption which is only acceptable in settings with low to moderate transmission level, as in [Champagne et al., 2022]. Relapses are modelled to occur at a constant rate, based on [White et al., 2016] which relied on [White et al., 2014], although the underlying mechanism could be represented with increased details [Mehra et al., 2022]. This compartmental model also relies on various homogeneity assumptions, as differences in infectiousness or susceptibility related to severity levels or age groups are not represented. Finally, the back-calculation methodology for baseline calibration relies on the steady state of the model, nonetheless other statistical methods could be used instead when temporal data is available [Chatzilena et al., 2019].

Despite these limitations, this model provides a useful additional tool to support country-specific decision making in the choice of interventions to deploy in various areas. Thanks to its analytical foundations and its simplicity of implementation, this model can support decision making on malaria strategies in a rapid and transparent manner.

## Data Availability

The computer code utilized in this manuscript is available at: https://github.com/SwissTPH/VivaxModelR.

## Author contributions

**Clara Champagne:** Conceptualization, Methodology, Software, Formal Analysis, Investigation, Writing Original Draft. **Maximilian Gerhards:** Conceptualization, Methodology, Formal Analysis, Investigation, Writing - Original Draft. **Justin Lana:** Conceptualization, Investigation, Writing - Review & Editing. **Arnaud Le Menach:** Conceptualization, Writing - Review & Editing, Project Administration, Funding Acquisition. **Emilie Pothin** Conceptualization, Methodology, Writing - Review & Editing, Project Administration, Funding Acquisition.

## Acknowledgements

The authors want to thank Gonché Danesh for her help with the *TiPS* package [Danesh et al., 2020, 2021], Roland Goers for code review, Jeanne Lemant for support in the parameterisation of IRS and Nakul Chitnis and Aatreyee M. Das for discussions on reactive case detection and for sharing their manuscript ahead of publication. Code development was performed on the infrastructure provided by sciCORE (http:////scicore.unibas.ch/), scientific computing center at University of Basel.

This work was supported, in whole or in part, by the Bill & Melinda Gates Foundation [OPP1109772/INV-008108]. Under the grant conditions of the Foundation, a Creative Commons Attribution 4.0 Generic License has already been assigned to the Author Accepted Manuscript version that might arise from this submission.

## A Including delay in treatment access: rationale for the chosen model

The model without treatment delays [Champagne et al., 2022] is defined by the following system of ordinary differential equations:

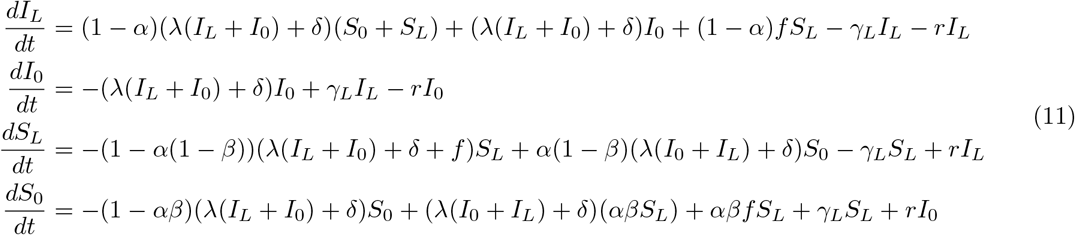

In this model, the effect of treatment is instantaneous, however, in reality individuals do not get treated instantaneously. Therefore, we aim to modify the model to include some time delay between infection and clearance of the parasite for treated individuals. We define *I* := 1 − *S*_*L*_ + *S*_0_ the proportion of individuals with blood-stage infections and will focus on the case with *β* = 1 (perfect radical cure for treated individuals).

Following the approach of Arino and Van den Driessche [Arino and van den Driessche, 2006] (vaccination model), we have the following equation for *I*:

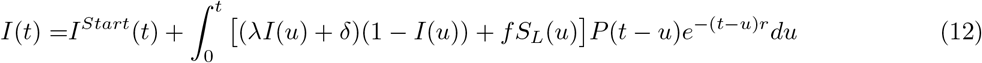

where *P* (*t* − *u*) is the proportion of those who were infected at time *u* and have not been effectively treated at time *t*. Two choices for the function *P* are explored in the following sections: the first one represents a delay with constant duration and the second one a delay with geometric duration.

### A.1 Delay with constant duration: using delay differential equations

If effective treatment is applied to a proportion *α* after a fixed time *σ*−^1^, the function *P* is a step function

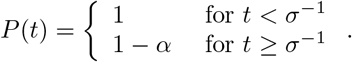

Inserting in the equation (12) we obtain the following equation:

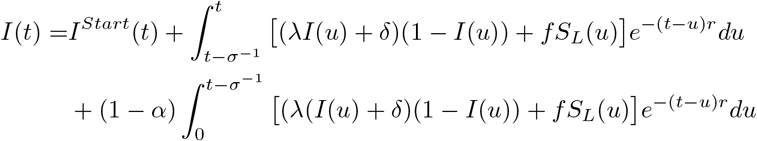

Differentiation yields the following delay differential equation:

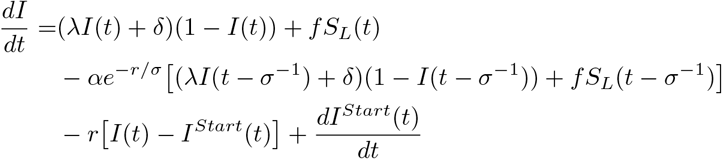

### A.2 Delay with exponential waiting time: using ordinary differential equations

If effective treatment is applied to a proportion *α* after a exponentially distributed waiting time with mean *σ*−^1^, the function *P* is of the form

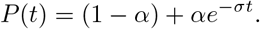

If we assume this treatment scheme to apply to those starting being infected, the equation 12 reduces to

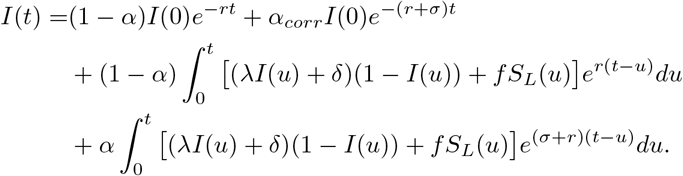

Differentiation yields

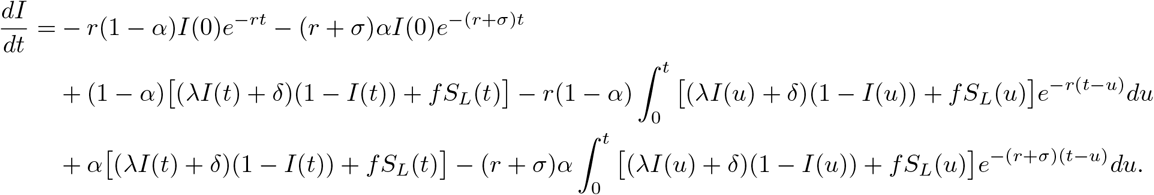

We can simplify this by separating those *α* that get effective treatment from those 1 − *α* that don’t. If we consider this separation not at the time the treatment is effective but at the time of infection, this defines two separate classes *U* for those that don’t have access to treatment and *T* for those that have access to treatment into which *α* of the newly infected get sorted. Of these, 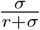 actually get effective treatment, after an average waiting time of 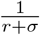, accounting for the possibility to recover before receiving the effective treatment. Then, we have a system of integral equations

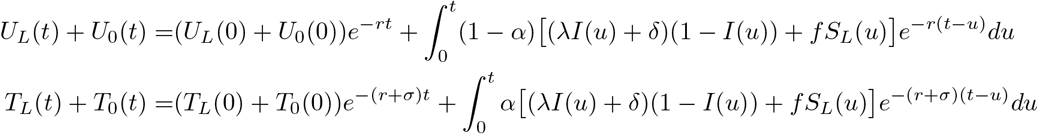

which when differentiated yield

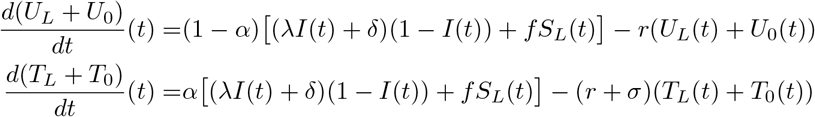

and therefore to

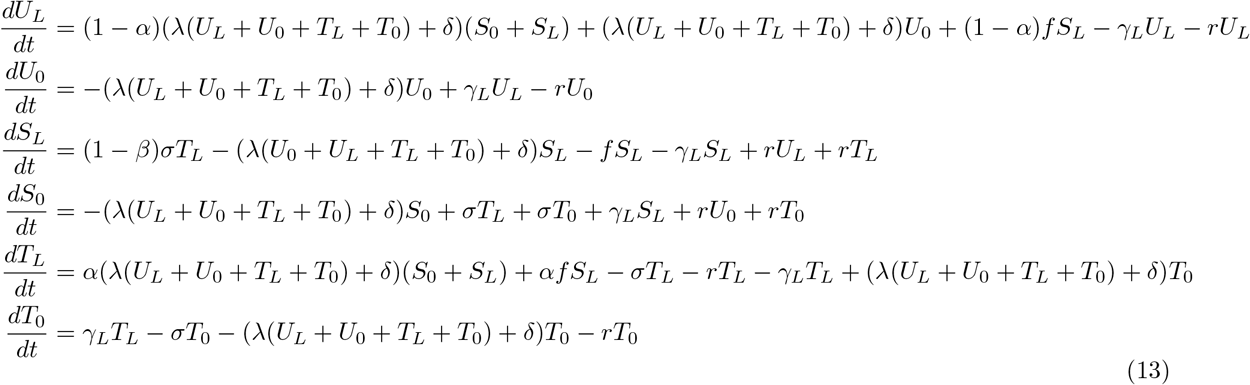

which is equivalent to the model with delayed treatment in the case *β* = 1.

As a final remark, one can note that taking the limit *σ* → ∞ we get the initial model with instantantaneous treatment (11), while letting *σ* = 0 we get the model (11) without treatment, as is to be expected.

It is worth noting that the actual probability of getting treated in both scenarios is not *α* (since some infected will already have recovered by the end of the delay), but 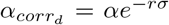 in the model with with constant delay and 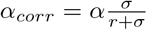 in the model with geometrically distributed delay. These numbers are also the proportions by which the average length of infection is reduced by including treatment.

These results are not easily generalized to the case where *β* < 1, due to the complexity of the integral equations. Nonetheless, the model (13) was extended to the case with imperfect radical cure (*β* < 1) leading to model (1) presented in the main text.

## B Back-calculation of the transmission rate in the model with delayed treatment

### B.1 Model equilibrium

Let 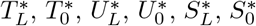, and *I*\* be the equilibrium proportions.

At the equilibrium, we have the equations

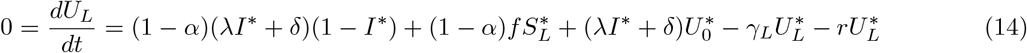

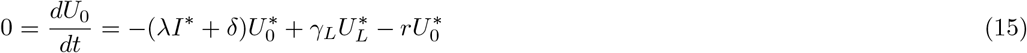

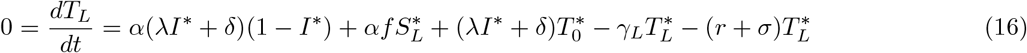

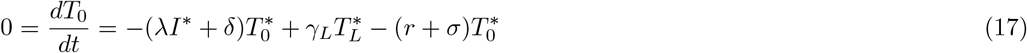

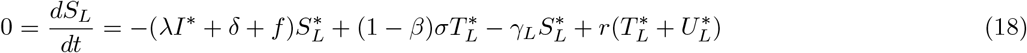

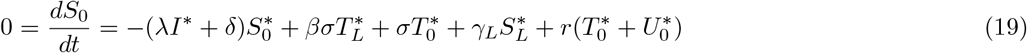

By adding equations (16) and (17) we obtain the additional equation:

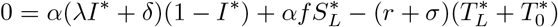

Likewise, by adding equations (14) and (15) we obtain the further equation:

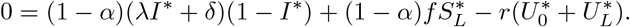

Hence we find the relations

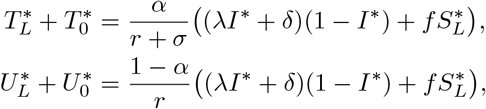

which can be combined to get

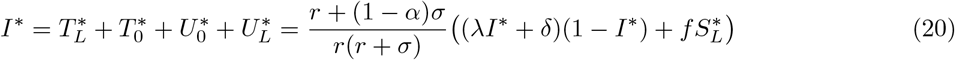

and

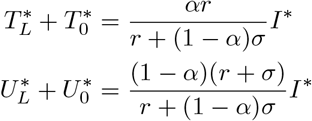

We define the observed incidence 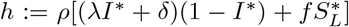 as the rate of observed newly arising blood-stage infections, where *ρ* is a reporting rate. Starting from equation (20), we can calculate *I*\* from observed quantities and model parameters as:

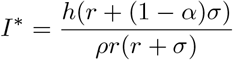

As *r* > 0 is necessary for the denominator to not be zero and is verified in all biologically plausible cases, we will continue with this assumption throughout the rest of the paper. If on the other hand *h* = 0, we have *I*\* = 0. Being in the disease-free equilibrium makes it impossible to derive *λ*. Because of this, we will also make the further assumption *h* > 0. It is worth noting that in this model, as opposed to that without treatment delay, *α* = 1 is not ruled out.

The proportion *p* of imported cases is defined such that *ph* := *ρδ*(1 − *I*\*) represents the imported cases and 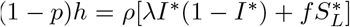 the locally acquired cases. Therefore, *δ* can be derived from observed quantities and model parameters exactly as in the model without delay:

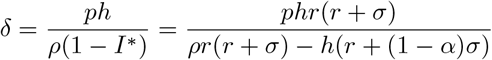

We then rely on the equilibrium relationships to calculate *λ* based on observed incidence *h* and the other model parameters.

We start by solving equation (17) for 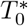:

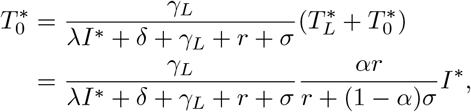

from which we arrive at the equation

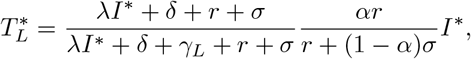

(neither of the denominators is zero as we assumed *r* > 0).

Likewise, we solve equation (15) for 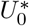:

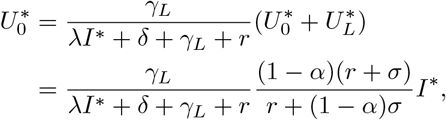

from which we arrive at the equation

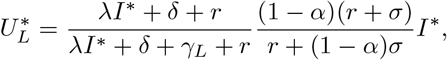

(again, neither of the denominators is zero). Solving (18) for 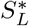 gives:

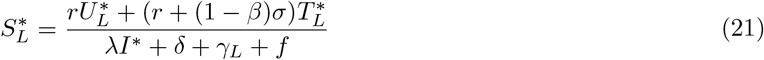

(the denominator cannot be 0, since *λI*\* = *δ* = *f* = 0 would imply *h* = 0).

Plugging both the equations for 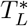 and 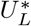 into (21) yields:

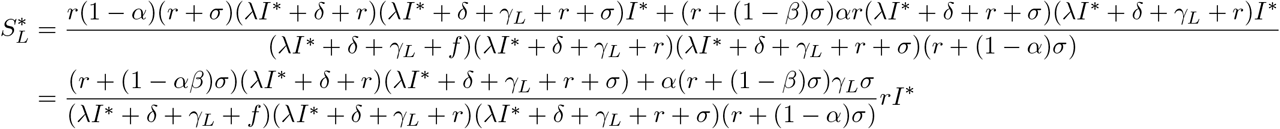

We can simplify further by using the identity 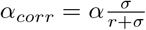:

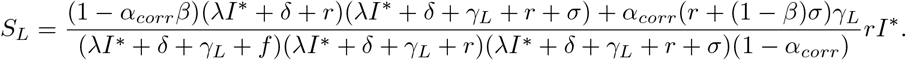

### B.2 Polynomial equation

Now, plugging this into (20) and multiplying by (*λI*\* + *δ* + *γ*_*L*_ + *f*)(*λI*\* + *δ* + *γ*_*L*_ + *r*)(*λI*\* + *δ* + *γ*_*L*_ + *r* + *σ*) we obtain:

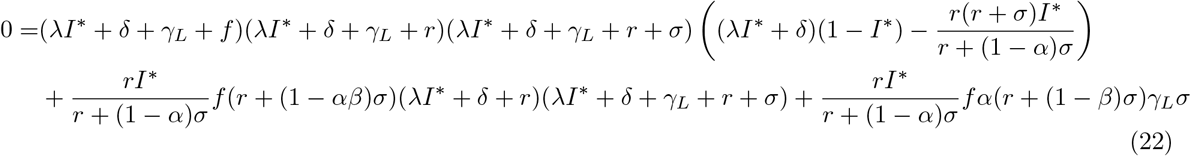

or, equivalently,

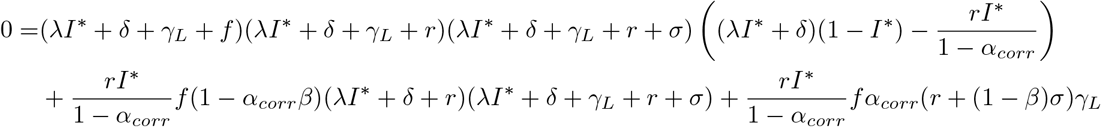

Rearranging the terms by powers of *λ*, we get the equation:

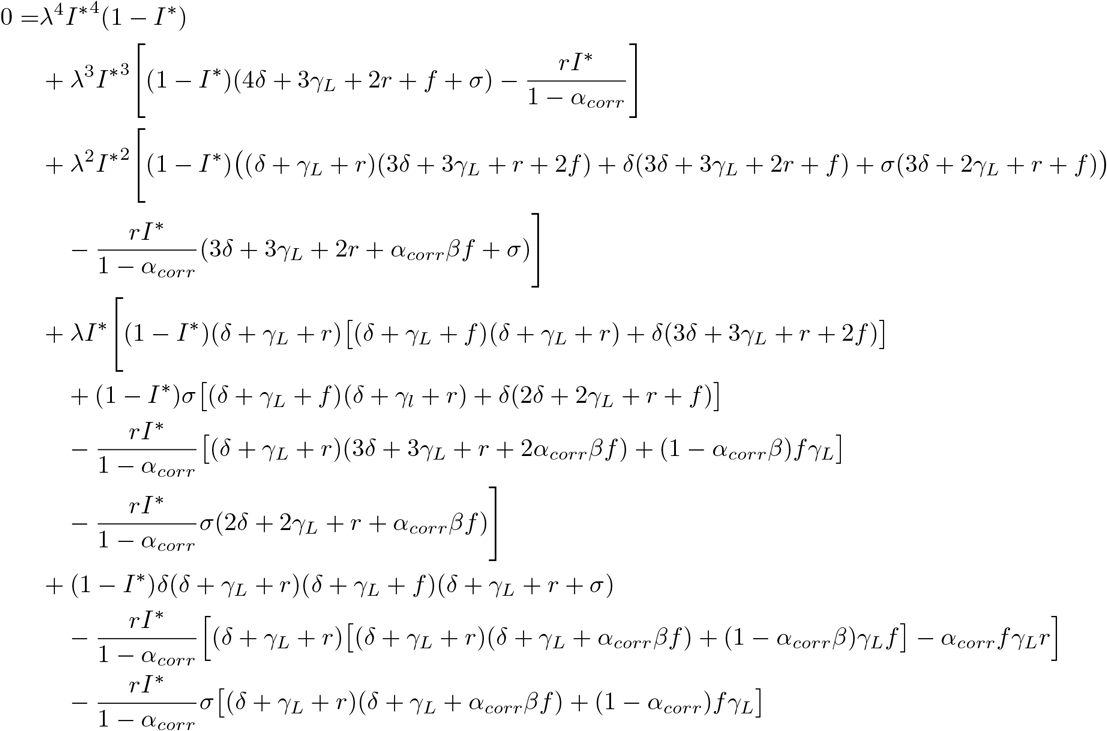

As long the assumptions *r* > 0 and *h* > 0 are met, multiplication by the denominator of 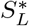 is an equivalent transformation, so any non-negative root of this polynomial is a solution of the system of equilibrium equations.

It can be noted that the equation in the setting without delay can be derived from this one by dividing by *σ* and taking the limit *σ* → ∞ (corresponding to a delay decreasing to 0).

We get the same qualitative result as in the setting without delayed treatment:

#### Theorem B.1.

*If h* > 0 *and r* > 0, *the function*

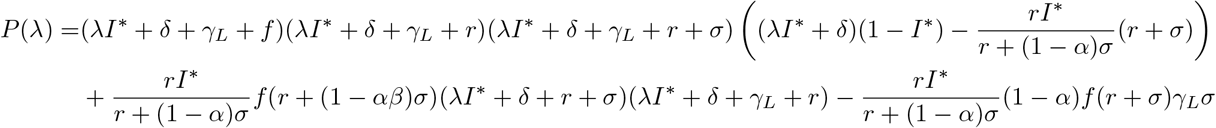

*has at most one positive real root*.

*It has two non-negative real roots (i*.*e. one of them is* 0*) only if αβσ* = *γ*_*L*_ = *δ* = 0 *(corresponding to an equilibrium of relapses and recoveries without liver-stage clearance)*.

Thanks to this result, it is possible to solve the polynomial equation numerically in order to back-calculate the parameter *λ* required to reproduce the reported data, assuming that the model is at equilibirum.

*Proof of Theorem B*.*1*. Before beginning the proof, we point to the fact that *h* > 0 and *r* > 0 imply 0 < *I*\* < 1.

We start with the case *f* = 0 like in the model without delay **?**.

From the shape of equation 22 one can easily see that the four roots in that case are

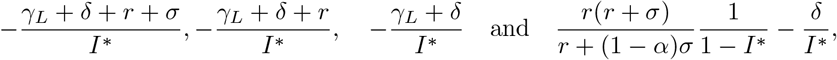

ordered from smallest to largest. Only the last two of them may be non-negative, and the first of these only in the case *γ*_*L*_ = *δ* = 0, and in that case, from *λ* = *δ* = *f* = 0 it follows *h* = 0, contradicting our assumption, so this is no solution.

Now we turn to the case *f* > 0.

To make the dependence of *P* on *f* visible, let us denote it *P*_*f*_. Then,

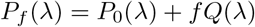

with

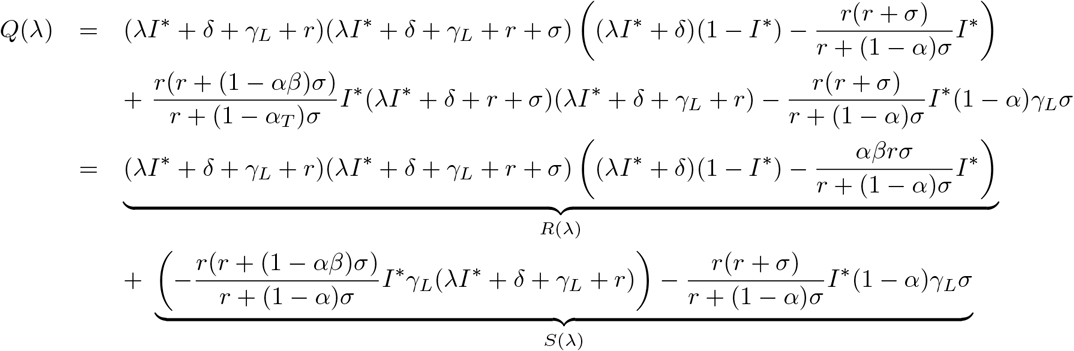

It is easily seen that *P*_0_ + *f R* is a polynomial of degree 4 with positive leading coefficient and two roots at 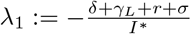 and 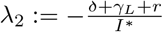.

Let us now first consider the case *γ*_*L*_ > 0. In that case we also see that

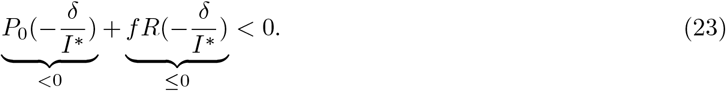

As lim_*λ*→∞_ *P*_0_(*λ*) + *f R*(*λ*) = ∞, from the intermediate value theorem it follows that *P*_0_ + *f R* has a root *λ*_4_ to the right of 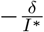. Since every simple root is accompanied with a sign change, the last root *λ*_3_ has to be to the left of 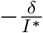. In particular, *λ, λ* and *λ* are all negative.

We will now prove that by adding 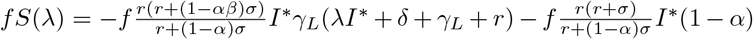, we will not get a non-negative root:

It is easily seen that *f S*(*λ*) < 0 for all *λ* ≥ 0, so

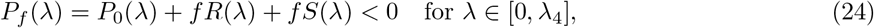

implying that there cannot be a root in that interval.

On the other hand, it is a known fact that the inflection points of a polynomial of degree 4 lie between the smallest and the largest root. This implies that *P*_0_ + *f R* is strictly convex in the interval]*λ*_4_, ∞[. Since *P*_*f*_ = *P*_0_ + *f R* + *f S* has the same second derivative, it is also strictly convex in that interval. This, combined with the fact that *P*_*f*_ (*λ*_4_) = *f S*(*λ*_4_) < 0, implies that there is only one root of *P*_*f*_ in that interval. This finishes the proof in the case *γ*_*L*_ > 0.

Now we turn to the case *γ*_*L*_ = 0. Then, *P*_*f*_ = *P*_0_ + *f R*.

We again know the two roots 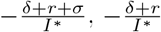 and we see that

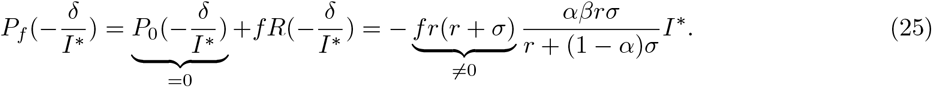

If this term is 0 (which is the case if and only if *αβσ* = 0), we know that 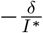 is also a root of *P*_*f*_. This root is only non-negative if *δ* = 0, which is the case of endless relapses and recoveries without liver-stage clearance.

Otherwise, the term must be negative. Then again, since every simple root is accompanied with a sign change, we know that there must be exactly one root to the right of 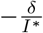.

## C Back-calculation of the transmission rate in the models with RCD

### C.1 Model with RCD and without delayed treatment

In this section exclusively, we use the notations *I* = *I*_*L*_ + *I*_0_ and 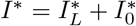.

In the absence of delays in treatment, the model with RCD is described by the following equations:

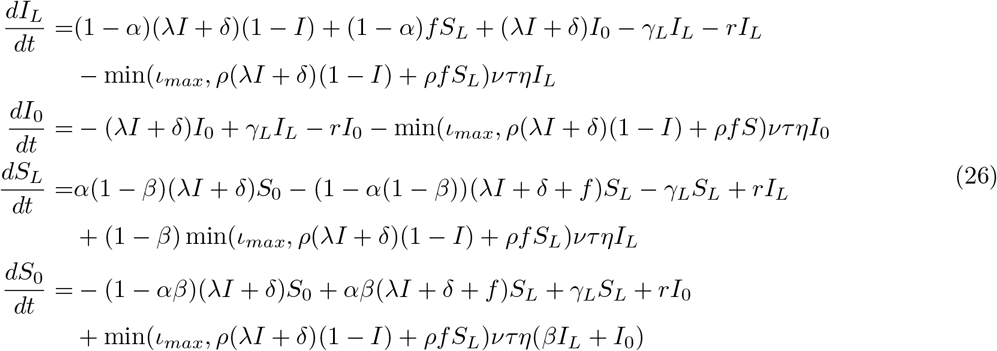

In the case of *ι*_*max*_*ντη* = 0, this reduces to the model without treatment delays in Champagne et al. [2022].

If there is no importation (*δ* = 0) and *τ* is fixed (or at least bounded), all of the ‘RCD terms’ are of the order *O*(*I*^2^) for *I* → 0, so the *R*_*C*_ value is equal to the *R*_*C*_ without any RCD, similarly to the model with treatment delays (4).

#### C.1.1 Model equilibrium

At equilibrium, adding the equations for *I*_*L*_ and *I*_0_ in (26), we get

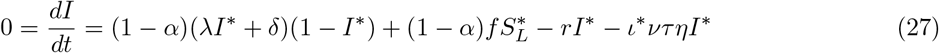

where 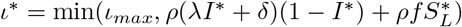.

Following the same steps as in the model without RCD [Champagne et al., 2022], we find

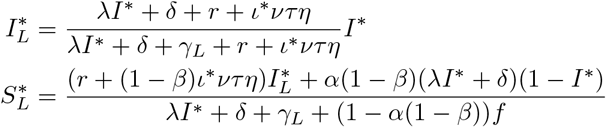

#### C.1.2 Polynomial equation

Solving these equations for *λ* gives the following polynomial equation:

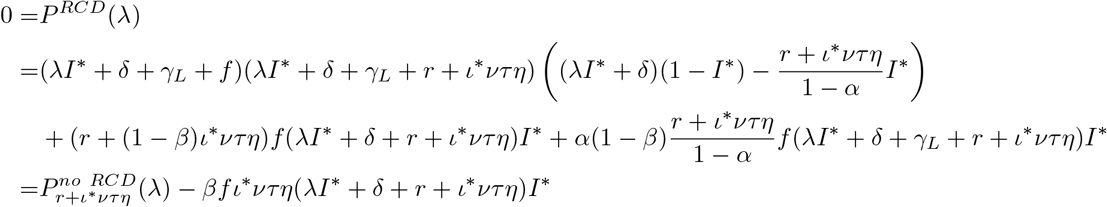

where 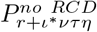 is the corresponding polynomial in the setting without RCD [Champagne et al., 2022], but with *r* replaced by *r* + *ι*\**ντη*. This equation can be used to back-calculate the transmission rate *λ* from observable quantities. The same formulae as in section 2.2.1 can be used to calculate *ι*\*, *I*\* and *δ*, using *I* = *I*_*L*_ + *I*_0_ and *α* instead of *α*_*corr*_.

The singularity of the solution can be shown similarly to the case without RCD [Champagne et al., 2022].

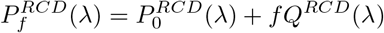

with

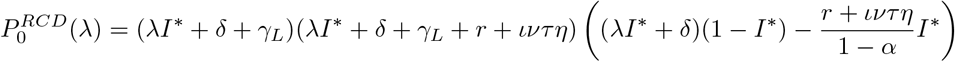

and

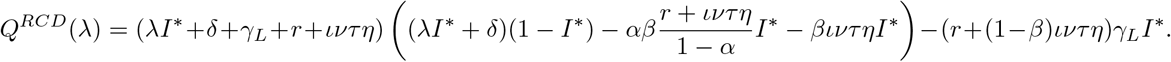

As expected, at equilibrium, for *β* = 0, adding the RCD term is equivalent to changing *r* to *r* +*ι*\**ντη* (but note that *ι*\* depends on the equilibrium states of the model *I*\* and 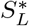). As *β* approaches 1, the difference between the two models increases.

### C.2 Back-calculation of the transmission rate in the model with delayed treatment and RCD (no referral)

In this section, additional results on the model (4) are provided.

#### C.2.1 Relation to other models

First, the model represented by (4) is related to other models in the following way:

- In the limit of *σ* → ∞, the model is equivalent to the non-delay RCD model (26).
- There are some more equivalences if *ι* is fixed instead of capped:

- If *β* = 0 and *σ* > *ιντη*, the model is equivalent to the model with delayed treatment and no RCD (1) with *r* replaced with *r* + *ιντη* and *σ* replaced with *σ* − *ιντη*.
- If *β* = 0 and *σ* < *ιντη*, the model is instead equivalent to the model with delayed treatment and no RCD (1) with *r* replaced with *r* + *σ, σ* replaced with *ιντη* − *σ, α* replaced with 1 − *α* and *T*_*L*_ and *T*_0_ swapped with *U*_*L*_ and *U*_0_, respectively.
- If *β* = 0 and *σ* = *ιντη*, the model is equivalent to the non-delay vivax model [Champagne et al., 2022] with *r* replaced with *r* + *σ, α* set to 0 and *T*_*L*_ and *T*_0_ added to *U*_*L*_ and *U*_0_, respectively.
- If *σ* = 0, the model is equivalent to the model with delayed treatment and no RCD (1) with *σ* replaced with *ιντη, α* replaced with 1 − *α* and *T*_*L*_ and *T*_0_ swapped with *U*_*L*_ and *U*_0_, respectively.

Such correspondences are used in the R package to test the correctness of the model implementation.

#### C.2.2 Model equilibrium

At equilibrium, adding the equations for *U*_*L*_ and *U*_0_ in (4), we get

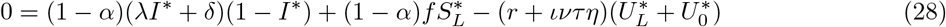

and similarly by adding the equations for *T*_*L*_ and *T*_0_ in (4),

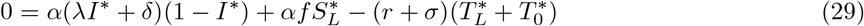

Finally, by adding both of these, we find

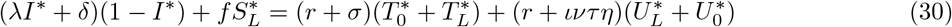

which can be inserted into either (28) or (29) to get

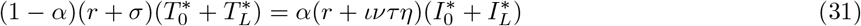

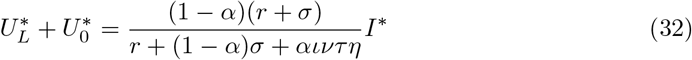

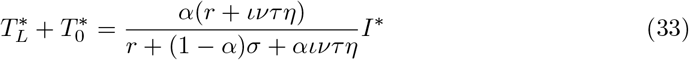

From the equation for *U*_0_ in (4), we obtain

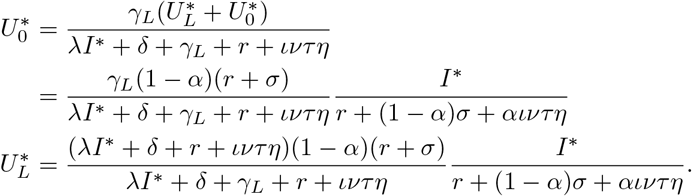

From the equation for *T*_0_ in (4), we obtain

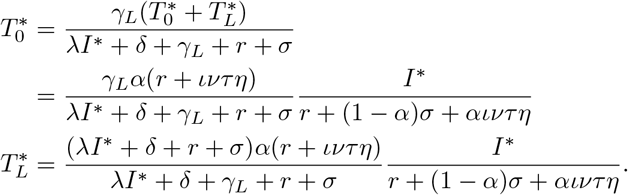

Inserting these results into the equation for *S*_*L*_ in (4), yields

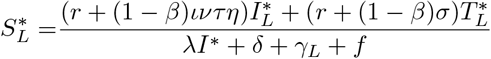

#### C.2.3 Polynomial equation

Plugging this into (30) yields

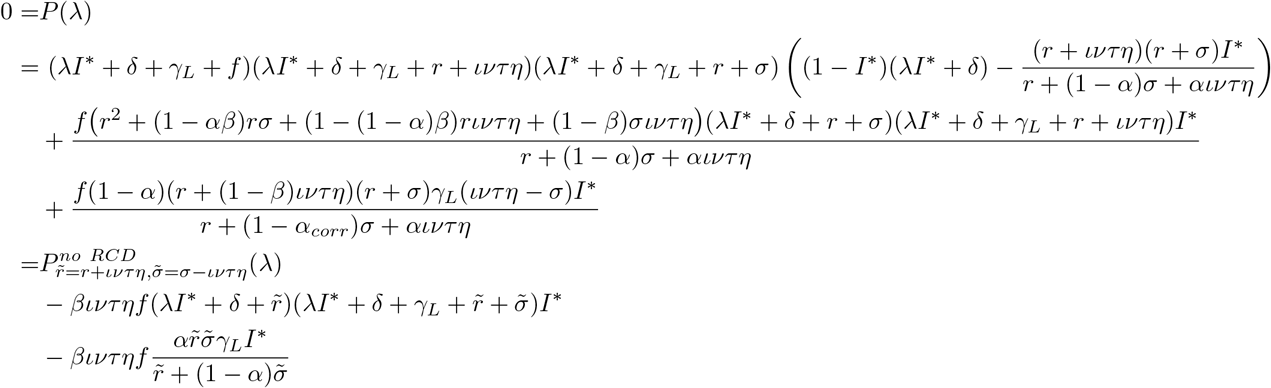

This equation can be used to back-calculate the transmission rate *λ* from observable quantities.

### C.3 Model with delayed treatment and RCD, including referral to health facility for cases detected via RCD

The model with delay in treatment and referral of reactively detected cases can be formulated as follows (noting *I* = *U*_*L*_ + *U*_0_ + *T*_*L*_ + *T*_0_):

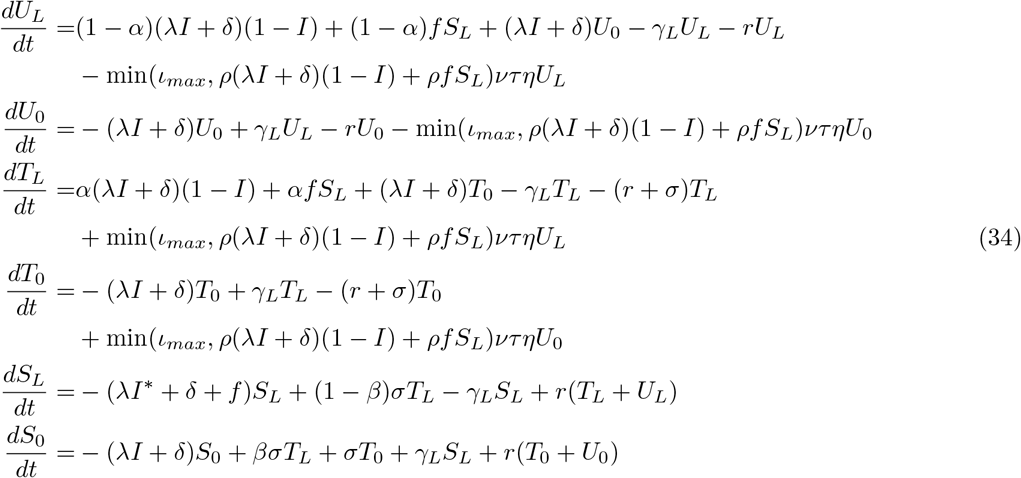

Similarly to model (4), if there is no importation (*δ* = 0) and *τ* is fixed (or at least bounded), all of the ‘RCD terms’ are of the order *O*(*I*^2^) for *I* → 0, so the *R*_*C*_ values are the same and equal to the *R*_*C*_ without any RCD.

#### C.3.1 Relation to other models

Again, we can make some connections to other models:

- If *ιντη* = 0, this model is the same as the model with delayed treatment and no RCD (1).
- In the limit of *σ* → ∞, the model is equivalent to the non-delay RCD vivax model (26).
- If *σ* = 0, the model is equivalent to the non-delay vivax model without either treatment or RCD [Champagne et al., 2022] with *T*_*L*_ and *T*_0_ added to *U*_*L*_ and *U*_0_, respectively.

#### C.3.2 Model equilibrium

Adding the equations for *U*_*L*_ and *U*_0_ in (34), we get

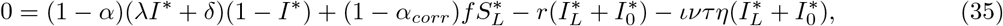

and similarly by adding the equations for *T*_*L*_ and *T*_0_ in (34),

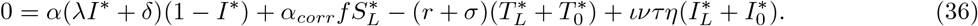

Finally, by adding both of these, we find

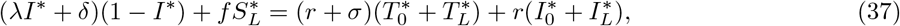

which can be inserted into either (35) or (36) to get

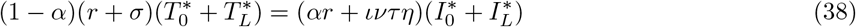

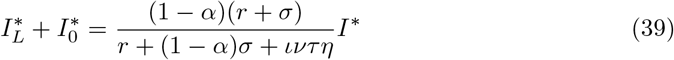

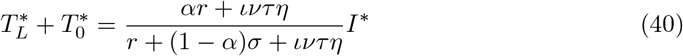

From the equation for *U*_0_ in (34), we obtain

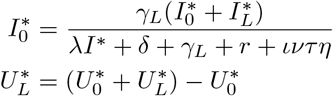

From the equation for *T*_0_ in (34), we obtain

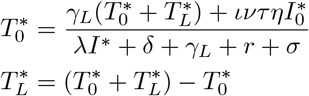

Inserting these results into the equation for *S*_*L*_ in (34) yields

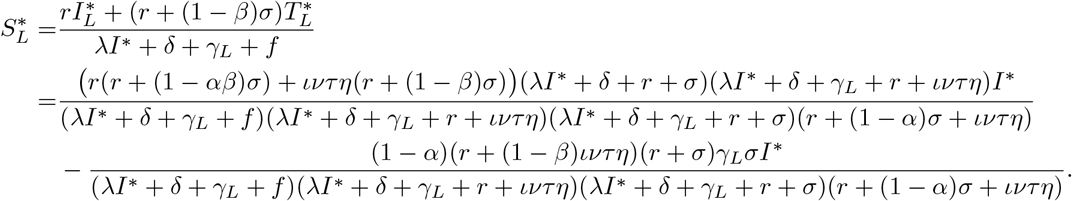

#### C.3.3 Polynomial equation

Plugging this into (37) yields

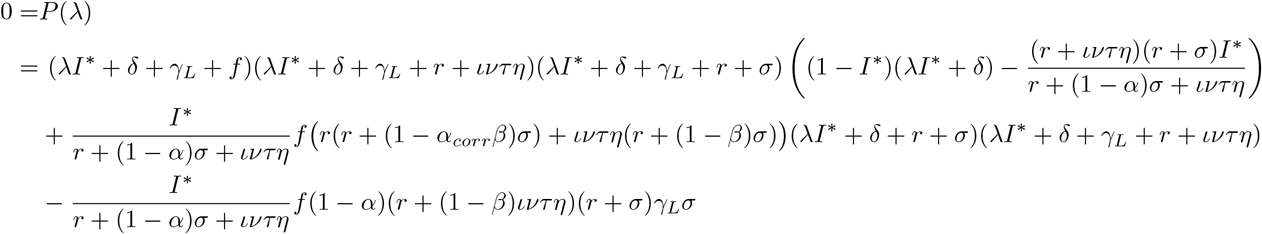

This equation can be used to back-calculate the transmission rate *λ* from observable quantities. The same formulae as in section 2.2.1 can be used to calculate *ι*\*, *I*\* and *δ*.

## D. Description of the other models including MDA

### D.1 No treatment delays, no RCD

The model is represented by the following ODE system:

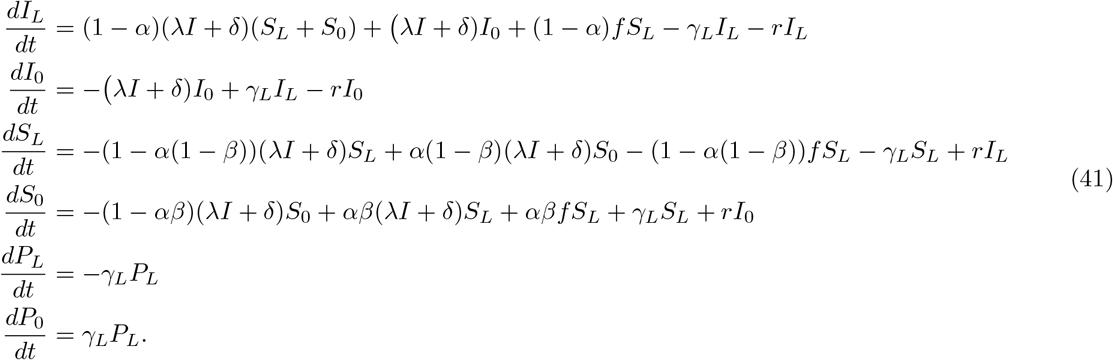

starting with *P*_*L*_(0) = *P*_0_(0) = 0.

At time *t*^*MDA*^, we take the new values

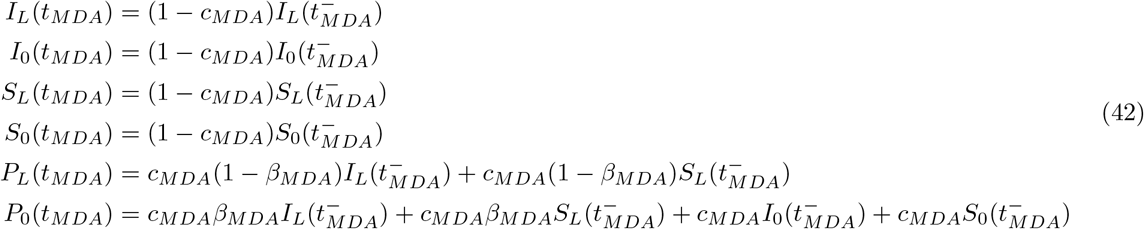

and at time *t*_*MDA*_ + *p*_*MDA*_, we take the values

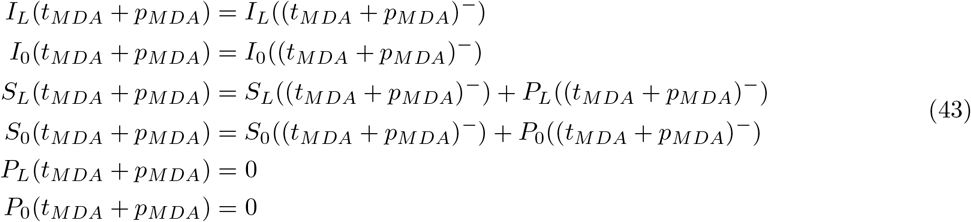

and continue the simulation from there.

### D.2 No treatment delays, with RCD

The model is represented by the following ODE system:

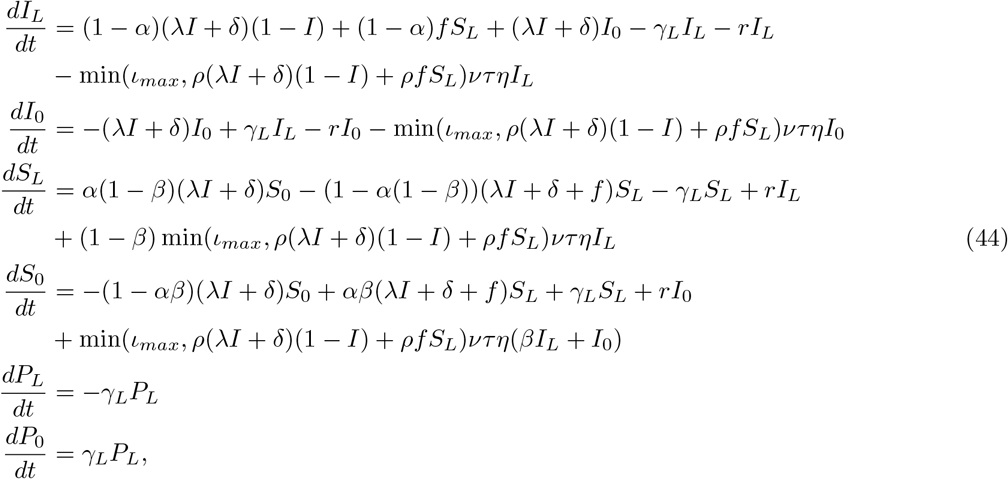

starting with *P*_*L*_(0) = *P*_0_(0) = 0.

At time *t*_*MDA*_, we take the new values

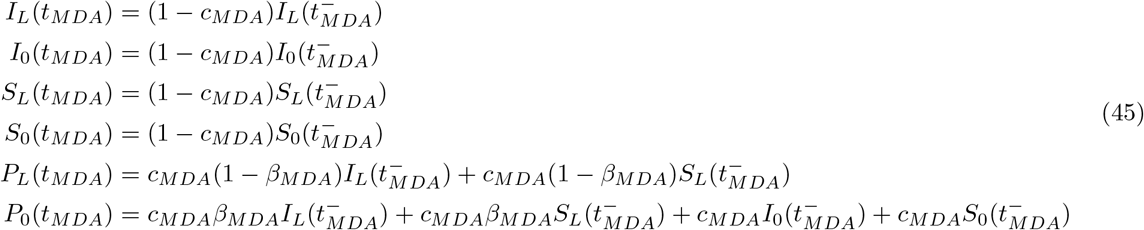

and at time *t*_*MDA*_ + *p*_*MDA*_, we take the values

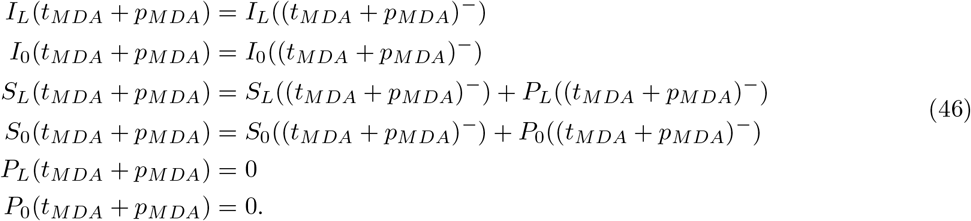

### D.3 RCD without referral to health facilities

The model is represented by the following ODE system:

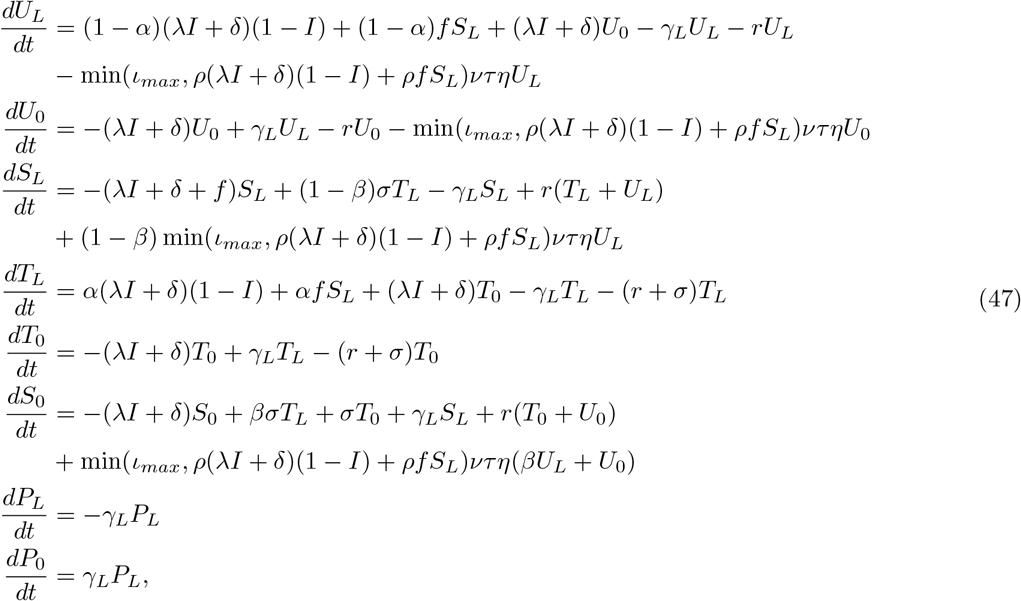

starting with *P*_*L*_(0) = *P*_0_(0) = 0.

At time *t*_*MDA*_, we take the new values

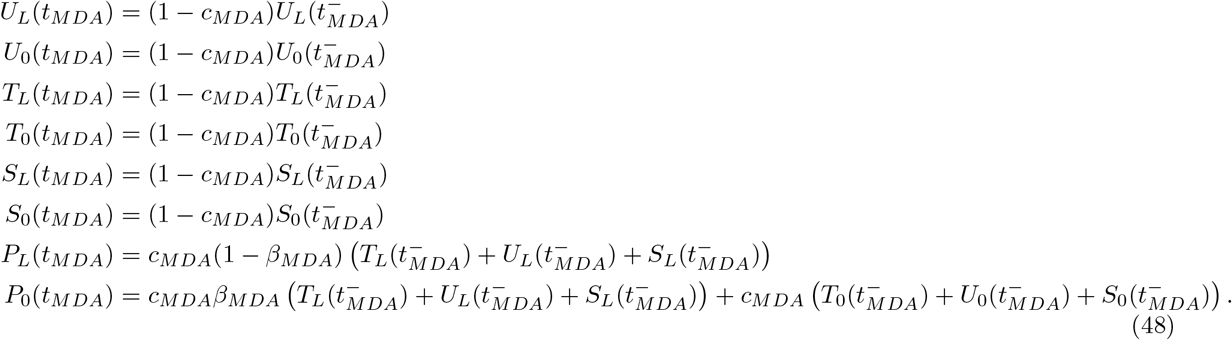

At last, at time *t*_*MDA*_ + *p*_*MDA*_ we take the new values

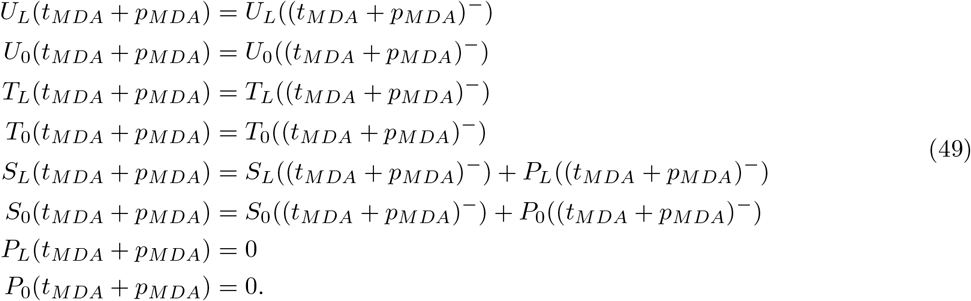

### D.4 RCD with referral to health facilities

The model is represented by the following ODE system:

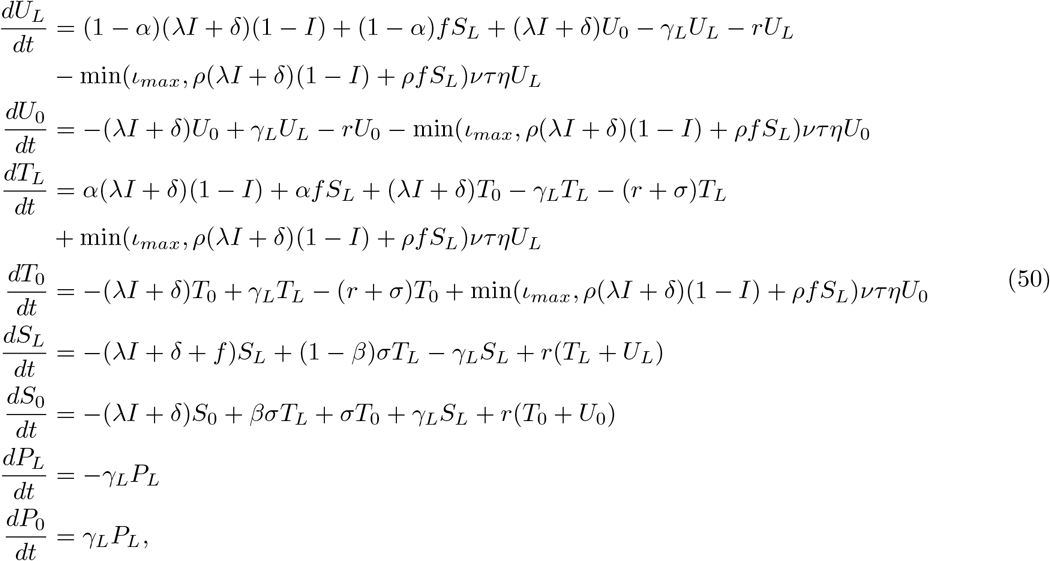

starting with *P*_*L*_(0) = *P*_0_(0) = 0.

At time *t*_*MDA*_, we take the new values

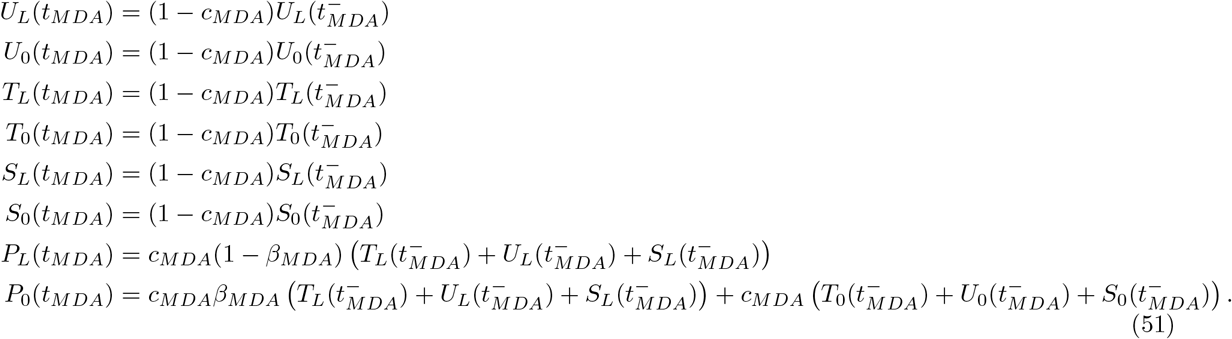

At last, at time *t*_*MDA*_ + *p*_*MDA*_, we take the new values

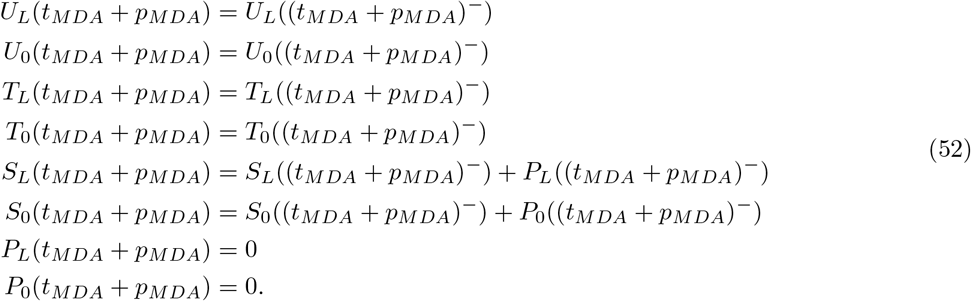

